# CD38^+^CD27^−^ on *Mtb*-specific CD4^+^ T cells distinguishes latent from active tuberculosis

**DOI:** 10.1101/2020.08.26.20180539

**Authors:** Muthya Pragun Acharya, Sai Pallavi Pradeep, Venkataramappa Srinivasa Murthy, Panduranga Chikkannaiah, Vivekanand Kambar, Satyanarayana Narayanashetty, Sharath Burugina Nagaraja, Niveditha, Raksha Yoganand, Vijaya Satchidanandam

## Abstract

**RATIONALE:** Early and accurate diagnosis followed by timely treatment are the key prerequisites to fight tuberculosis (TB) and reduce its global burden. Despite scientific advances, the rapid and correct diagnosis of both pulmonary and extrapulmonary tuberculosis remains a challenge due to traditional reliance on detection of the elusive bacilli. *Mycobacterium tuberculosis* (*Mtb*)-specific host immune activation and cytokine production has shown significant promise as an alternative means of detecting and distinguishing active disease from latent infection.

**OBJECTIVE:** Phenotypic characteristics of *Mtb*-specific cytokine-producing immune cell subsets were investigated and queried for their diagnostic ability in identifying active tuberculosis.

**METHODS:** Subjects belonging to the following groups were recruited – pulmonary, extrapulmonary, latent TB, cured TB, sick controls and healthy controls. Polychromatic flow cytometry was used to identify host immune biomarkers in an exploratory cohort comprising 56 subjects using peripheral blood mononuclear cells. Clinical performance of the identified biomarker was evaluated using whole blood in a blinded validation cohort comprising 165 individuals.

**FINDINGS:** Frequencies of *Mtb*-specific CD4^+^ T cells of the phenotype CD38^+^CD27^−^ clearly distinguished patients with active tuberculosis from individuals without the disease. CD38^+^CD27^−^CD4^+^ T cells secreting TNF-α upon stimulation with ESAT6/CFP10 peptides had the best diagnostic accuracy at a cut-off of 9.91% [exploratory: 96.67% specificity, 88.46% sensitivity; validation: 96.15% specificity, 90.16% sensitivity]. Additionally, this subset differentiated treatment-naive TB patients from individuals cured of TB following completion of anti-tuberculosis therapy.

**INTERPRETATION:** *Mtb*-specific CD38^+^CD27^−^TNF-α^+^CD4^+^ T cell subset is a robust biomarker for TB diagnosis and can determine cure.

**IMPACT OF THIS RESEARCH:** We identified and validated CD38^+^CD27^−^TNF-α^+^ as a robust biomarker with diagnostic accuracies >90% in both PBMCs and whole blood that can be translated into a reliable and cost-effective in vitro diagnostic test with ease. By not removing samples with insignificant immune response and instead classifying them as negative, our study represents a truly realistic assessment of the diagnostic accuracy of the identified biomarker in a clinical setting.

## INTRODUCTION

Tuberculosis which remains one of the world’s deadliest infectious diseases with significant morbidity and mortality affects roughly 10 million people and claims 1.5 million lives annually (1). In India alone, 2.4 million people were affected by TB in 2019 (2). Major hurdles in controlling TB include high number of latently infected individuals (LTBI) (approximately 23% of the world’s population, of which 5–15% can progress to active disease in their lifetime), delayed diagnosis and missed TB cases (̴ 3 million), the latter two contributing to TB transmission (1). A 2015 retrospective analysis of 3220 TB patients diagnosed between 2005 and 2011 found that 25.7% of them had prior hospital visits for respiratory illness within a 90-day window preceding their TB diagnosis (3).

Smear microscopy, the commonly used test for the diagnosis of pulmonary TB (PTB) has poor sensitivity (4). Even though nucleic acid amplification tests such as GeneXpert have shown higher sensitivity in smear/culture positive cases, they perform poorly against composite reference standards (5). The execution of liquid culture tests, the gold standard for TB diagnosis, despite its accuracy has high turnover time and is extremely demanding in resource constrained settings (6). The painful invasive procedures to obtain clinical samples combined with its paucibacillary nature makes diagnosis of extrapulmonary TB (EPTB) all the more challenging. Relative to sputum samples, GeneXpert performs poorly in detecting bacilli in body fluids and aspirates (5, 7). If all these challenges associated with diagnosing TB can be overcome, effective treatment can be commenced early, reducing transmission and enhancing outcomes. Thus, a rapid and accurate diagnostic test will contribute significantly to controlling and eradicating TB in accordance with the END-TB Strategy (8).

Host based surrogate markers have been explored in recent times as a novel avenue for TB diagnosis. Activation (CD38, HLA-DR, CD127), differentiation (CD27, KLRG1), homing (CCR4, CCR7) and proliferation (Ki-67) markers on *Mtb*-specific CD4^+^ T cells and their functional profiles (IFN-ɣ, TNF-α, IL-2, IL-10, IL-17) have been extensively explored to identify potential biomarkers for the diagnosis of TB (9–11). CD38, HLA-DR and Ki-67 on *Mtb*-specific CD4^+^IFN-ɣ^+^ cells had impressive performance in discriminating LTBI from both PTB and EPTB and also in monitoring treatment response and assessing cure (12–16).

The robust cytokine TNF-α whose production at significantly higher frequencies makes its detection efficient has also been explored as a TB biomarker in numerous studies (9). The CD38^+^CD27^−^ phenotype on CD4^+^ and on *Mtb*-specific CD4^+^IFN-ɣ^+^ T cells has recently been shown to be dominant in active tuberculosis (aTB) (17). However, it’s biomarker performance was not evaluated. Here, we explored the ability of TNF-α secreting *Mtb*-specific CD38^+^CD27^−^CD4^+^ T cells to distinguish active from latent TB using PBMCs and validated its diagnostic performance in whole blood to enable translation into a field–compatible in vitro diagnostic test.

## METHODS

### Ethics statement

This study was carried out in accordance with the Declaration of Helsinki. Ethical clearance and necessary approvals were obtained from the Institutional Review Board of Indian Institute of Science (IISc), Bengaluru (5-14032018) and Employees State Insurance Corporation Medical College & Post Graduate Institute of Medical Sciences & Research (ESIC MC & PGIMSR), Bengaluru (532/L/11/12/Ethics/ESICMC&PGIMSR/Estt. Vol..III).

### Study participants

245 participants were recruited at ESIC MC & PGIMSR between Jan 2019 and Dec 2019. Written informed consent was obtained from all participants for the collection of samples and further analyses. 4 – 8mL blood was collected in sodium heparin prior to treatment initiation and matched sputum samples were collected wherever possible. HIV+/HBV+ individuals were excluded from analysis. Recruitment of study participants has been outlined in Figure 1 and Table E1. Patient details have been summarised in Tables E2–4.

**Figure 1.**
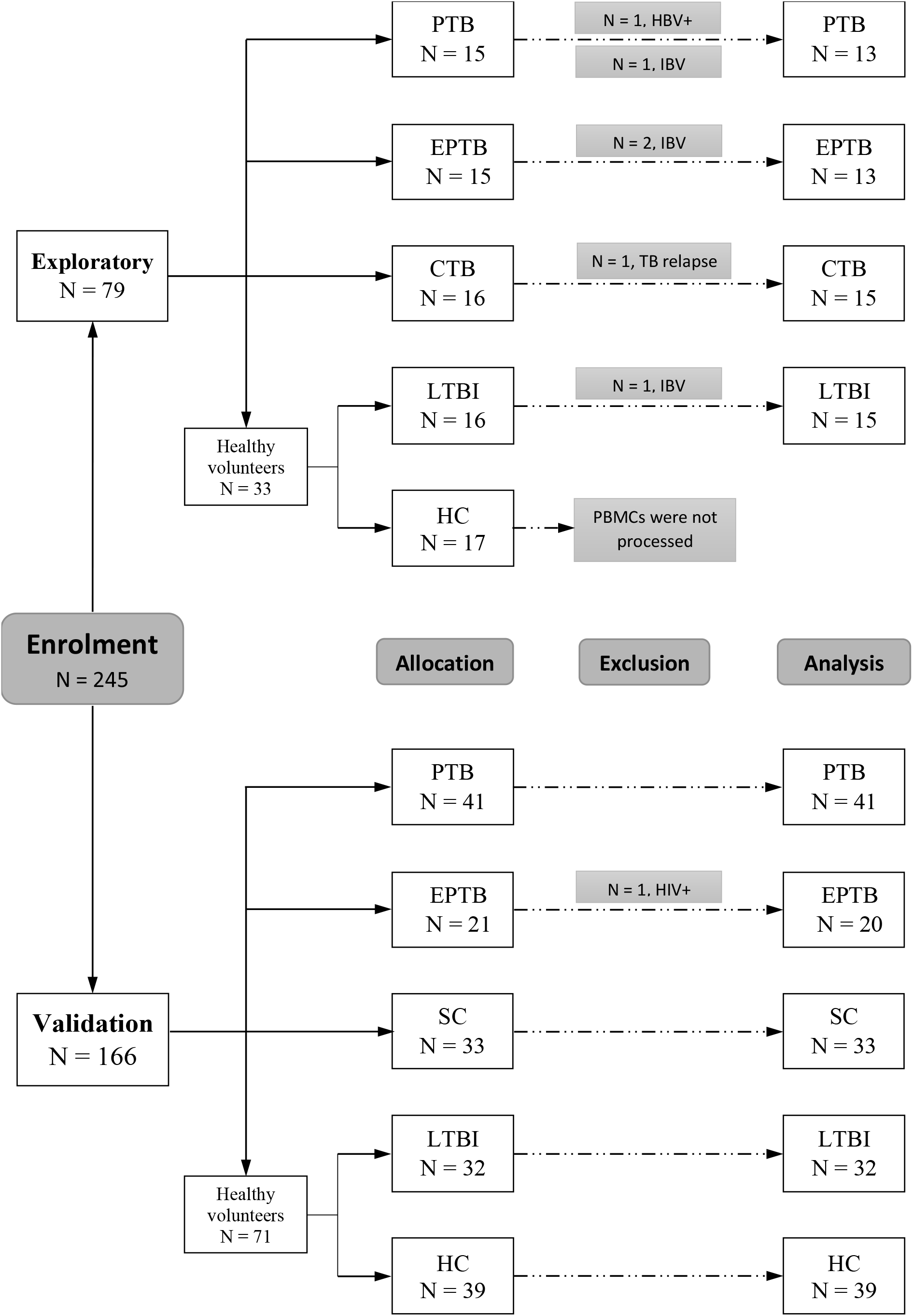
CONSORT flow diagram of recruited patients. 245 individuals were recruited for the study in two separate cohorts: exploratory and validation. The recruited individuals belonged to one of the following groups as indicated - pulmonary tuberculosis (PTB), extrapulmonary tuberculosis (EPTB), latent tuberculosis infection (LTBI), healthy controls (HC), cured tuberculosis (CTB) or sick controls (SC). HBV: Hepatitis B virus, HIV: Human immunodeficiency virus, IBV: insufficient blood volume.

### Antigens and antibodies

ESAT6/CFP10 peptides comprising 15mers with 11 amino acid overlap were procured from Synpeptide Co., Ltd (Shanghai, China). *Mtb*-cell wall antigen (*Mtb-*CW) was prepared as described previously (18, 19) and has been explained in detail in the online data supplement. The flow cytometry antibody panels used in the study are described in Table E5.

### PBMC isolation, stimulation and flow cytometry

PBMCs were isolated by density gradient centrifugation on Histopaque®-1077 (Sigma-Aldrich) and suspended in RPMI-1640 medium containing 10% FBS, 2mM glutamine, 100 IU/ml penicillin, and 100mg/ml streptomycin. One million PBMCs or 0.5mL whole blood were stimulated with ESAT6/CFP10 peptide pool or *Mtb-*CW antigen at 10µg/mL in presence of brefeldin A (10µg/mL, Biolegend) for 20h at 37°C, 5% CO_2_. Post incubation, PBMCs were first stained with Fixable Viability Stain 450 (FVS450, BD Biosciences) to discriminate live from dead cells, followed by appropriate surface markers. Cells were then fixed with 1% paraformaldehyde (10mins) and permeabilized with 0.1% saponin. Permeabilized cells were stained for appropriate intracellular markers and fixed with 0.5% paraformaldehyde before acquisition. For whole blood post antigen stimulation, RBCs were lysed and WBCs were fixed in a single lyse/fix step with 1mL of RBC lysis buffer (3% diethylene glycol, 1.875% - 2% formaldehyde and 0.5% - 0.75% methanol). The centrifuged cells were initially stained with FVS450 and then stained for both surface and intracellular markers in Buffer 2 (PerFix-nc kit; Beckman Coulter). The cells were then washed and resuspended in Buffer 3 (PerFix-nc kit; Beckman Coulter) prior to acquisition. Samples were acquired on Beckman Coulter DxFLEX flow cytometer and the data were analysed using CytExpert v2.0.

### Statistical analysis

Antigen specific responses were considered positive if they met the following criteria: (i) a frequency ≥ 0.05% in the stimulated tube and ≥ 0.03% over the unstimulated control for TNF-α; (ii) twice the frequency of that in the unstimulated control and ≥ 0.02% over and above it for IFN-ɣ; (iii) a frequency ≥ 0.02% over the unstimulated control for IL-2. Additionally, responses were considered significant only if at least 15 events were recorded. Statistical analyses were performed using GraphPad Prism 6.

## RESULTS

### Activated and differentiated CD4^+^ T cell phenotypes are more prevalent in aTB

PBMCs from 56 subjects – 13 PTB, 13 EPTB, 15 cured TB (CTB) and 15 LTBI (Figure 1, Table 1 and Tables E1–3) were stimulated with ESAT6/CFP10 and *Mtb-*CW antigens. Gating strategy is shown in Figure E1. TNF-α responses from CD4^+^ T cells in the four groups are summarised in Figure 2. Despite a significant difference between EPTB and CTB upon ESAT6/CFP10 stimulation (Figure 2A), TNF-α^+^CD4^+^ T cells could not differentiate aTB from non-TB groups (LTBI and CTB) in contrast to an earlier report (20). Surprisingly, 7 of the 15 individuals (46.67%) in the CTB group who had recently completed ATT had no significant CD4^+^TNF-α^+^ production upon ESAT6/CFP10 stimulation along with 2 of the 13 (15.39%) in EPTB group. One individual in CTB group did not have significant response to either ESAT6/CFP10 or *Mtb-*CW antigen.

**Table 1.**
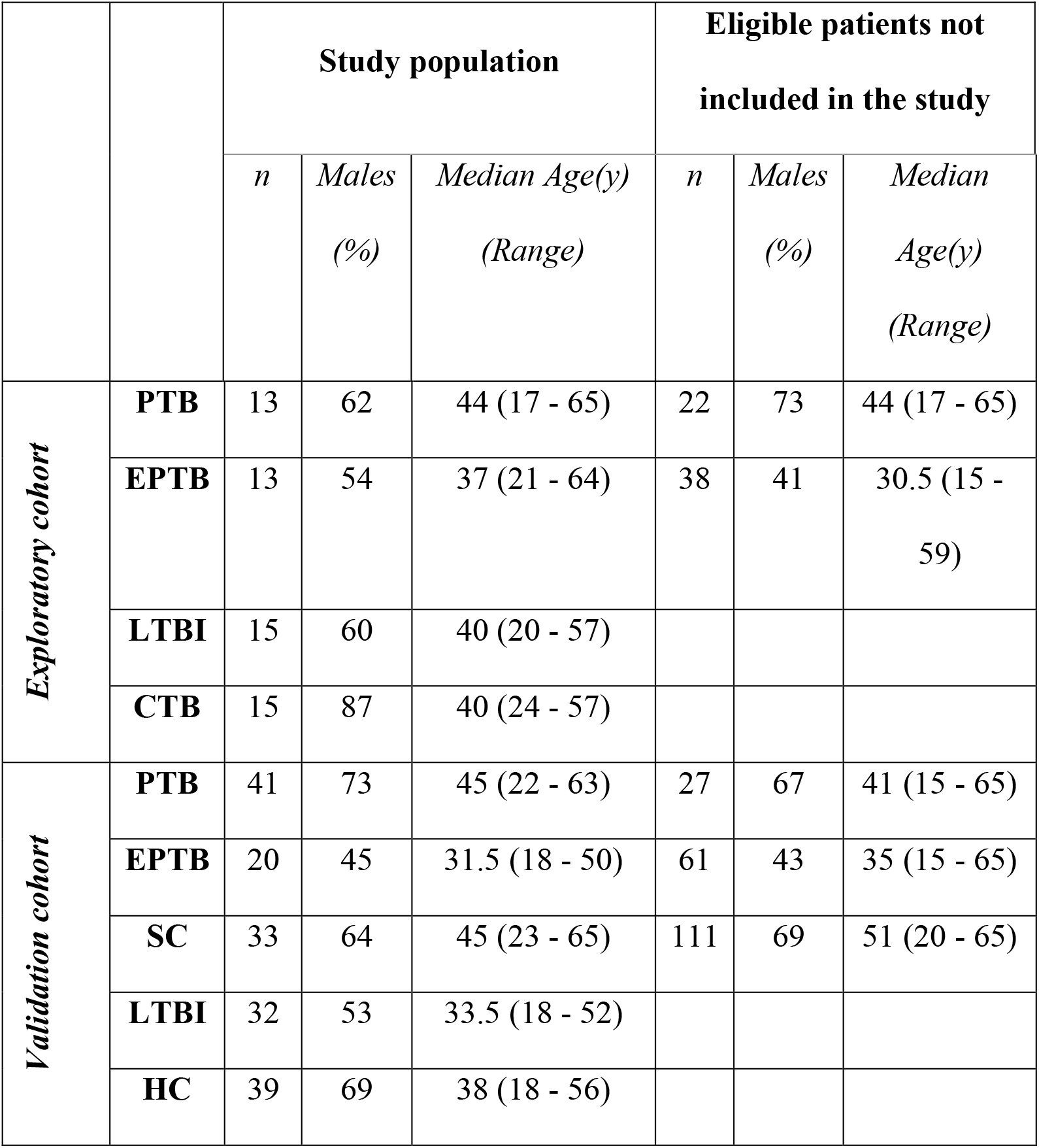
Characteristics of the study population. The characteristics have been compiled for patients whose samples were included for analysis. PTB: pulmonary tuberculosis, EPTB: extrapulmonary tuberculosis, LTBI: latent tuberculosis infection, HC: healthy controls, CTB: cured tuberculosis and SC: sick controls.

|  |  | Study population |  |  | Eligible patients not included in the study |  |  |
| --- | --- | --- | --- | --- | --- | --- | --- |
|  |  | *n* | *Males (%)* | *Median Age(y) (Range)* | *n* | *Males (%)* | *Median Age(y) (Range)* |
| *Exploratory cohort* | **PTB** | 13 | 62 | 44 (17 - 65) | 22 | 73 | 44 (17 - 65) |
|  | **EPTB** | 13 | 54 | 37 (21 - 64) | 38 | 41 | 30.5 (15 - 59) |
|  | **LTBI** | 15 | 60 | 40 (20 - 57) |  |  |  |
|  | **CTB** | 15 | 87 | 40 (24 - 57) |  |  |  |
| *Validation cohort* | **PTB** | 41 | 73 | 45 (22 - 63) | 27 | 67 | 41 (15 - 65) |
|  | **EPTB** | 20 | 45 | 31.5 (18 - 50) | 61 | 43 | 35 (15 - 65) |
|  | **SC** | 33 | 64 | 45 (23 - 65) | 111 | 69 | 51 (20 - 65) |
|  | **LTBI** | 32 | 53 | 33.5 (18 - 52) |  |  |  |
|  | **HC** | 39 | 69 | 38 (18 - 56) |  |  |  |

**Figure 2.**
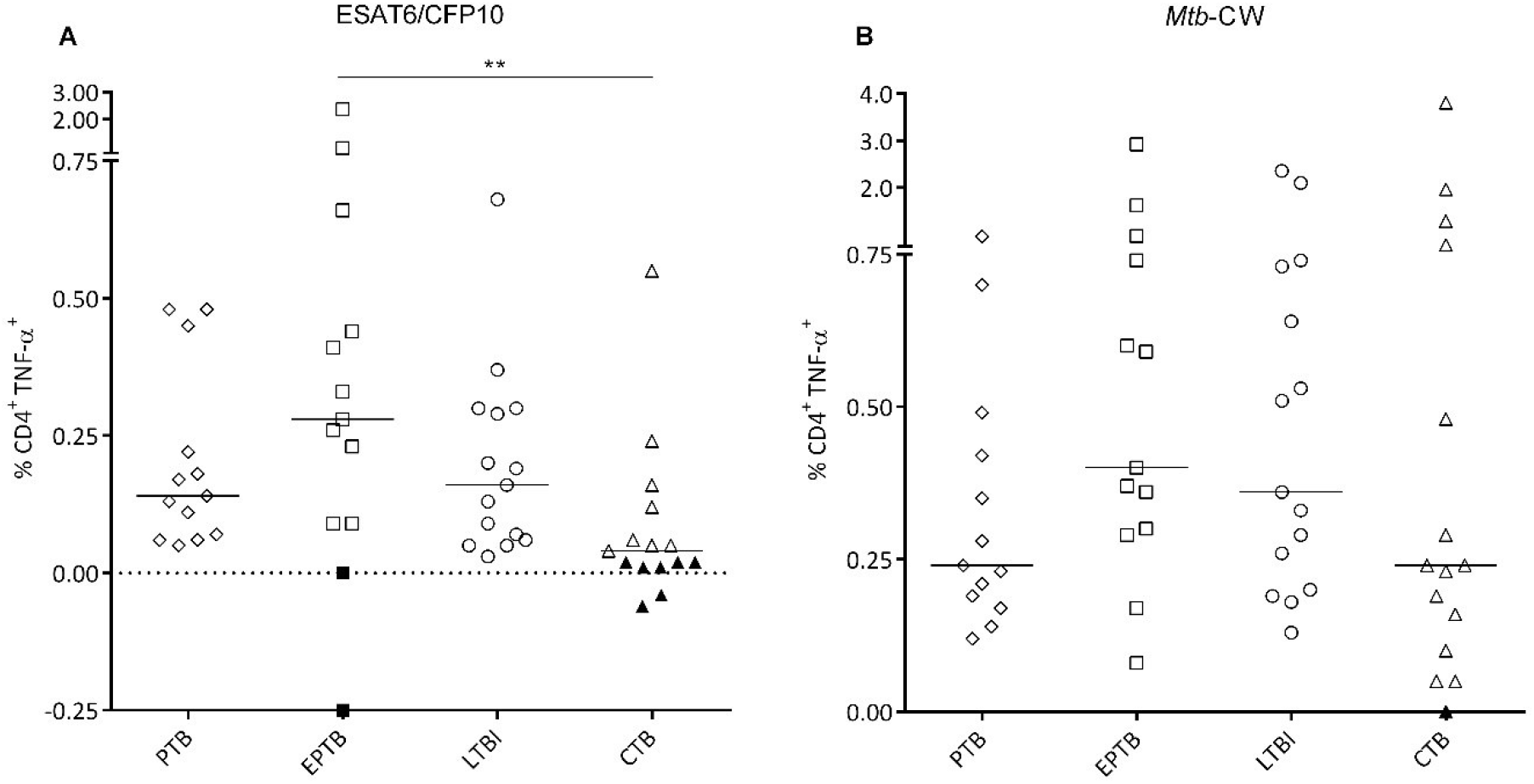
Frequencies of TNF-α secreting CD4^+^ T cells in the exploratory cohort. PBMCs from treatment naive PTB (n = 13) and EPTB (n =13) subjects, LTBI (n = 15) and CTB (n = 15) were stimulated with **(A)** ESAT6/CFP10 and **(B)** *Mtb-*CW antigens and analysed by flow cytometry. CD4^+^TNF-α^+^ frequencies are reported after subtracting the unstimulated response from the stimulated tubes. Kruskal-Wallis test followed by Dunn’s post-test was used to compare the groups: **p < 0.01. Filled symbols indicate samples with no-significant CD4^+^TNF-α^+^ response. Bars represent medians.

Expression of CD38^+^, CD27^−^, CD38^+^CD27^−^ and CD38^−^CD27^+^ was investigated on antigen specific TNF-α^+^CD4^+^ T cells by multiparameter flow cytometry (Figure E2). Frequencies of candidate biomarkers on *Mtb-*specific cells were normalised to zero in non-responders and were not removed from analyses. We observed that significantly higher proportions of *Mtb*-specific TNF-α^+^CD4^+^ T cells were phenotypically either CD38^+^, CD27^−^ or CD38^+^CD27^−^ in aTB compared to non-TB groups. In contrast, CD38^−^CD27^+^TNF-α^+^CD4^+^ T cells were significantly overrepresented in LTBI and CTB relative to aTB (Figure 3, A and B). In accordance with an earlier report (17), CD38^+^CD27^−^ frequencies on total CD4^+^ T cells were significantly higher in aTB while CD38^−^CD27^+^ was the predominant phenotype in non-TB groups (Figure 3C). Thus, these results indicate that activated and differentiated T cell phenotypes on total CD4^+^ and *Mtb*-specific TNF-α^+^CD4^+^ T cells are associated with active disease.

**Figure 3.**
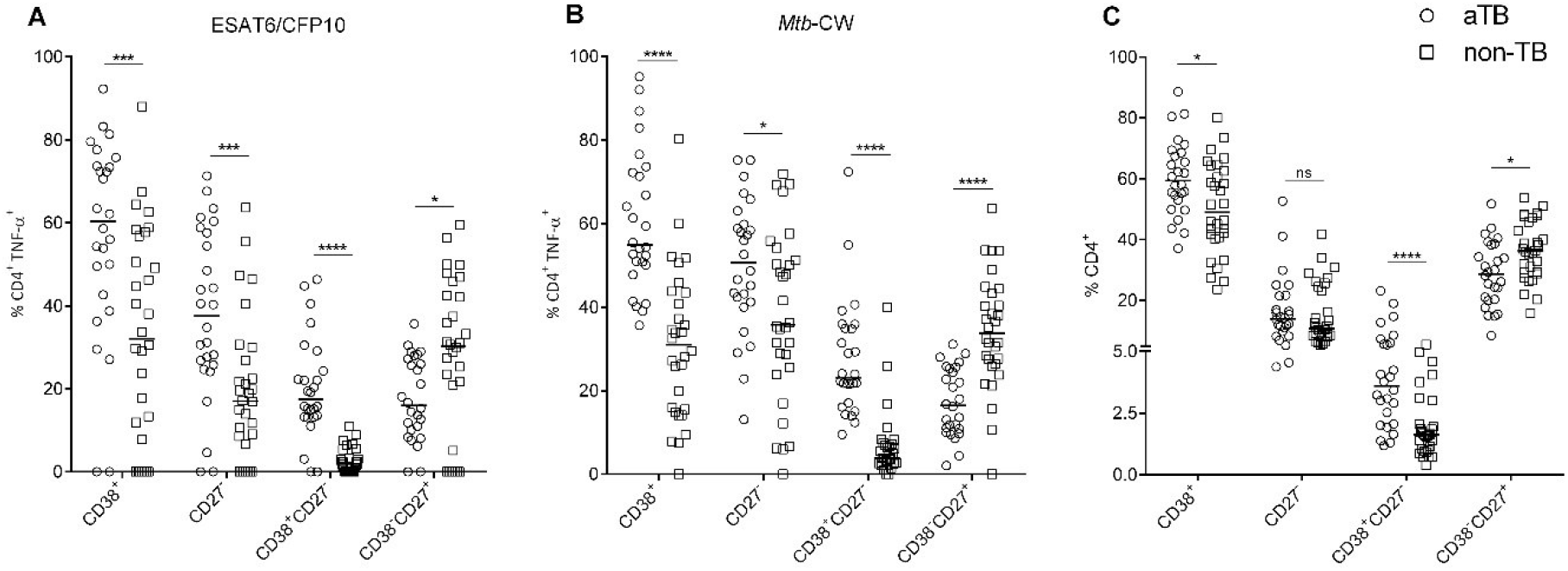
Expression of phenotypic markers on *Mtb*-specific TNF-α^+^CD4^+^ and total CD4^+^ T cells. CD38^+^, CD27^−^, CD38^+^CD27^−^ and CD38^−^CD27^+^ phenotypes on TNF-α^+^CD4^+^ T cells were analysed in PBMCs stimulated with **(A)** ESAT6/CFP10 and **(B)** *Mtb-*CW antigens from 26 individuals with aTB (circles) (13 PTB and 13 EPTB) and 30 individuals from non-TB groups (squares) (15 LTBI and 15 CTB). Frequencies of *Mtb*-specific TNF-α^+^CD4^+^ T cells with different phenotypic markers were normalised to 0% in samples with no significant TNF-α production. **(C)** Phenotypic characteristics on total CD4+ T cells in non-stimulated PBMCs. Data were analysed using Mann-Whitney *U* test: *p < 0.05; **p < 0.01; ***p < 0.001; ****p < 0.0001, ns: non-significant. Bars represent median values.

### CD38^+^CD27^−^TNF-α^+^CD4^+^ T cells can detect active tuberculosis

Receiver operating characteristic (ROC) analysis was performed to evaluate the predictive value of CD38^+^TNF-α^+^ (green), CD27^−^TNF-α^+^ (red), CD38^+^CD27^−^TNF-α^+^ (black) and CD38^−^CD27^+^TNF-α^+^ (blue) in correctly classifying the TB and the non-TB groups and AUC values were determined (Figure 4, A and B). CD38^+^CD27^−^TNF-α^+^ had the highest values for AUC with both ESAT6/CFP10 and *Mtb-*CW antigen stimulation - 0.93 and 0.94 respectively (Figure 4C). At a cut off value of 9.91%, CD4^+^CD38^+^CD27^−^ T cells secreting TNF-α accurately discriminated aTB from non-TB with a diagnostic accuracy of 92.86% with ESAT6/CFP10 stimulation (88.46% sensitivity and 96.67% specificity) and 91.07% with *Mtb-*CW stimulation (96.15% sensitivity and 86.67% specificity) (Figure 4D). When we excluded samples without significant responses from analysis as done in earlier reported studies, we obtained a much higher diagnostic accuracy of 95.74% (95.83% sensitivity and 95.65% specificity) with ESAT6/CFP10 stimulation (12, 21, 22). CD38^+^CD27^−^TNF-α^+^ frequencies in CTB were comparable with those in LTBI, suggesting that this population would also serve to monitor treatment responses and assess cure (Figure 4D). Thus, CD38^+^CD27^−^TNF-α^+^CD4^+^ T cells can accurately discriminate active TB irrespective of the disease site (both PTB and EPTB infections) from non-active states (LTBI and CTB).

**Figure 4.**
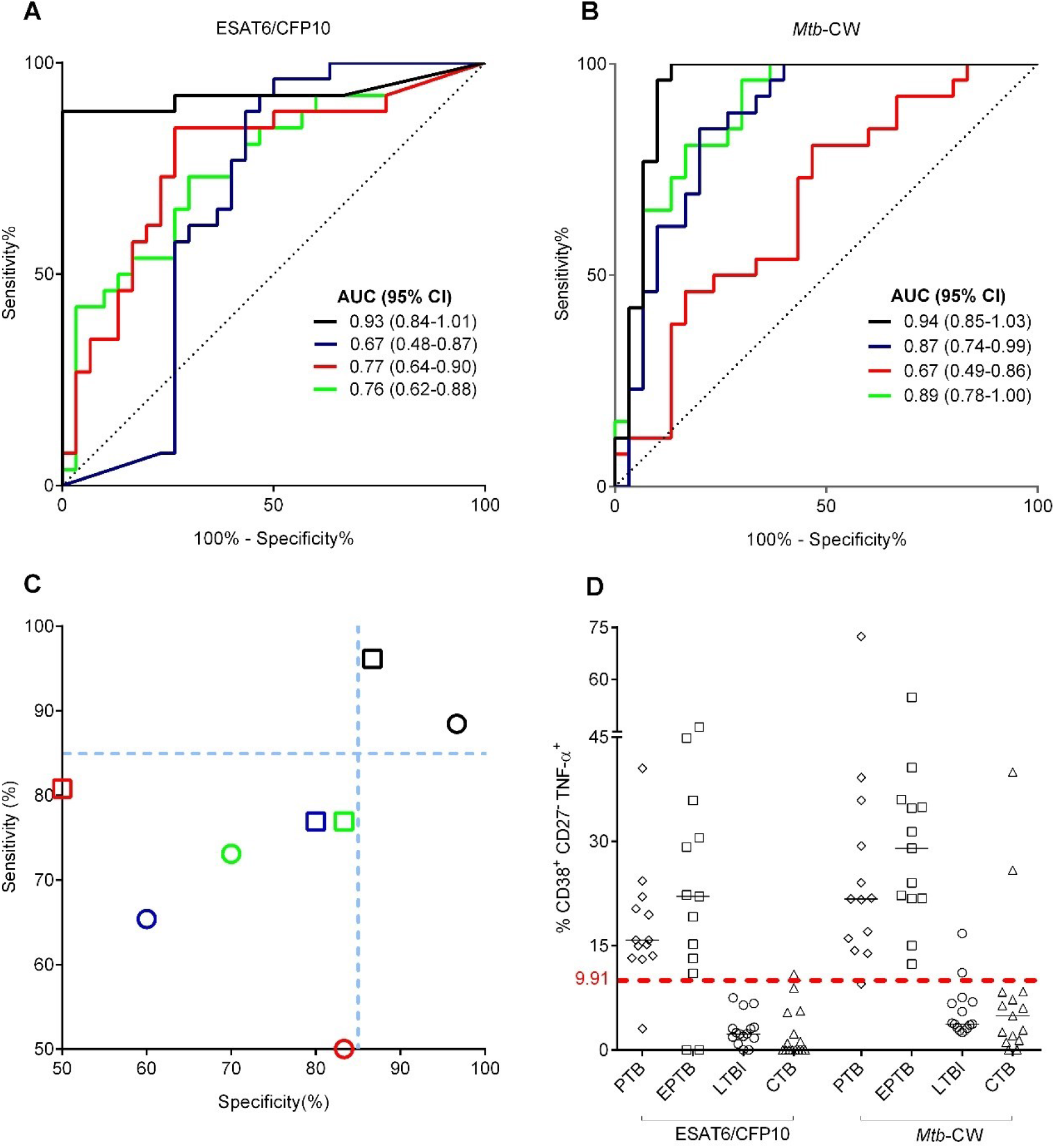
Evaluation of diagnostic performance of phenotypic markers on *Mtb*-specific TNF-α^+^CD4^+^ T cells in exploratory cohort. Receiver Operating Characteristics (ROC) curve analyses of CD38^+^ (green), CD27^−^ (red), CD38^+^CD27^−^ (black) and CD38^−^CD27^+^ (blue) phenotypes on *Mtb*-specific TNF-α^+^CD4^+^ T cells was used to determine their ability to discriminate aTB from non-TB groups upon **(A)** ESAT6/CFP10 and **(B)** *Mtb-*CW antigen stimulation. Dotted line represents the line of identity. AUC: area under the curve and CI: confidence interval **(C)** Diagnostic performance of the four biomarkers upon ESAT6/CFP10 (circle) and *Mtb-*CW antigen (square) stimulation. Dotted blue lines represent 85% sensitivity and specificity. **(D)** Frequencies of CD38^+^CD27^−^TNF-α^+^ in PBMCs stimulated with ESAT6/CFP10 and *Mtb-*CW antigen from individuals in the following categories: PTB (n = 13), EPTB (n = 13), LTBI (n = 15) and CTB (n = 15). CD38^+^CD27^−^TNF-α^+^ frequencies in samples with insignificant CD4+TNF-α+ response were normalised to 0%. The red dashed line represents the discriminatory cut-off value of 9.91%. Bars represent medians.

### Evaluation of predictive value of CD38^+^CD27^−^TNF-α^+^ in an independent blinded cohort

In order to validate the diagnostic performance of the candidate biomarker CD38^+^CD27^−^TNF-α^+^CD4^+^ T cells, we resorted to the use of whole blood instead of PBMCs in a blinded cohort with the aim to enhance the utility of the test for field settings. The tedious and expertise-requiring process of PBMC isolation is challenging to conduct in resource limited clinical settings with high TB burden. In contrast, use of whole blood is economical, time saving, technically simple and amenable for translation into an in vitro diagnostic test. 165 subjects belonging to either PTB, EPTB, LTBI, healthy controls (HC) or sick controls (SC) were recruited as described in Figure 1 and Tables E1-4. Whole blood was stimulated with both ESAT6/CFP10 and *Mtb-*CW antigen followed by flow cytometry analyses as mentioned in Methods. Frequencies of CD38^+^CD27^−^TNF-α^+^ in 48 and 14 individuals upon ESAT6/CFP10 and *Mtb-*CW stimulation respectively, where no significant CD4^+^TNF-α^+^ production was observed were normalised to zero. Using the predetermined cut off value, we observed that following ESAT6/CFP10 stimulation, 59 individuals had CD38^+^CD27^−^TNF-α^+^ frequencies above 9.91% and 106 individuals below it (Figure 5A). With *Mtb-*CW stimulation, 61 individuals had CD38^+^CD27^−^TNF-α^+^ frequencies above 9.91% and 104 individuals below it (Figure 5B). When unblinded, we observed that ESAT6/CFP10-specific CD38^+^CD27^−^TNF-α^+^ frequencies had correctly classified 55 of 61 aTB patients (36 of 41 PTB and 19 of 20 EPTB) and 100 of 104 non-TB individuals (31 of 32 LTBI, 30 of 33 SCs and 39 of 39 HCs) (Figure 5C and Table 2). *Mtb-*CW-specific CD38^+^CD27^−^TNF-α^+^ frequencies correctly classified 49 of 61 aTB patients (30 of 41 PTB and 19 of 20 EPTB) and 92 of 104 non-TB individuals (31 of 32 LTBI, 26 of 33 SCs and 35 of 39 HCs) (Figure 5D and Table E6). Thus, CD38^+^CD27^−^TNF-α^+^ frequencies correctly distinguished TB from non-TB with an accuracy of 93.94% with ESAT6/CFP10 stimulation (90.16% sensitivity and 96.15% specificity) and 85.45% with *Mtb-*CW stimulation (80.33% sensitivity and 88.46% specificity). However, *Mtb-*CW-specific CD38^+^CD27^−^TNF-α^+^ had a better accuracy of 87.88% (88.52% sensitivity and 87.50% specificity) at a cut off of 7.89% instead of 9.91% (Table E6). We then sought to render the test cost effective by confining the flow cytometry parameters to a single laser (5-parameter – CD3, CD4, CD27, CD38, TNF-α) by excluding the viability dye (FVS450) from analyses. The diagnostic accuracy was found to be 93.94% with ESAT6/CFP10 stimulation (91.80% sensitivity and 95.19% specificity) and 89.09% with *Mtb-*CW stimulation (88.52% sensitivity and 89.42% specificity) under these conditions (data not shown). Median frequencies of dead lymphocytes in the stimulated tubes were low (1.92%; Range – 0.90% to 9.50%) and ignoring the viability dye did not significantly alter the diagnostic performance.

**Table 2.**
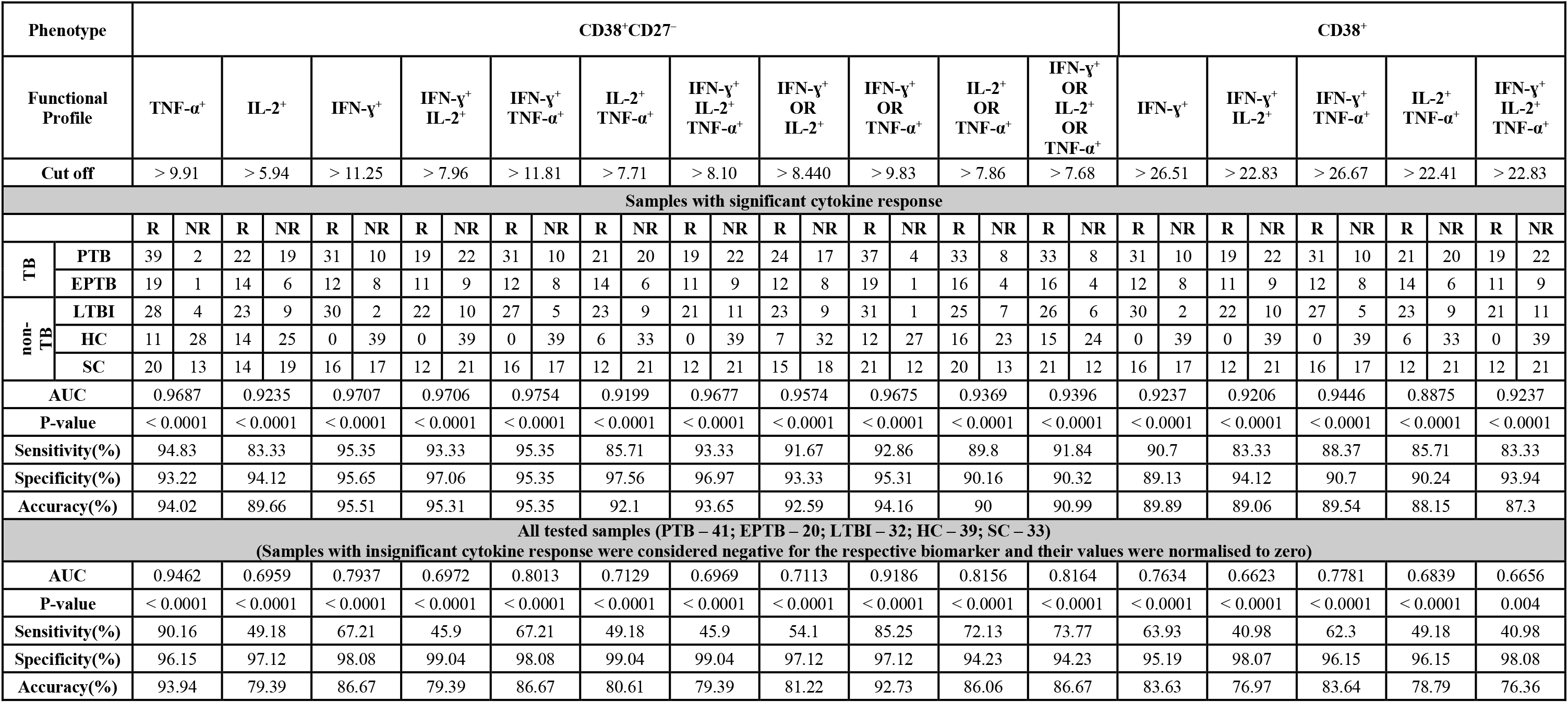
Biomarker performance of CD38^+^CD27^−^ and CD38^+^ within mono- and polyfunctional CD4^+^ T cells. Diagnostic potentials of several biomarkers with AUC values > 0.875 in ESAT6/CFP10 stimulated whole blood have been summarised. Number of samples with significant responses for each cytokine or their combination have been listed. For AND combinations, response was considered significant if all the cytokines involved in the combination were independently significant. For OR combinations, criteria for significant responses were same as that for TNF-α. To counter the problem of insignificant cytokine responses in whole blood, OR combinations were evaluated to check if antibodies for 2 or more cytokines could be used in the same detector channel. Mann Witney *U* test was used to compare the TB (PTB and EPTB) and non-TB (LTBI, HC and SC) groups. AUC: area under the curve.

**Figure 5.**
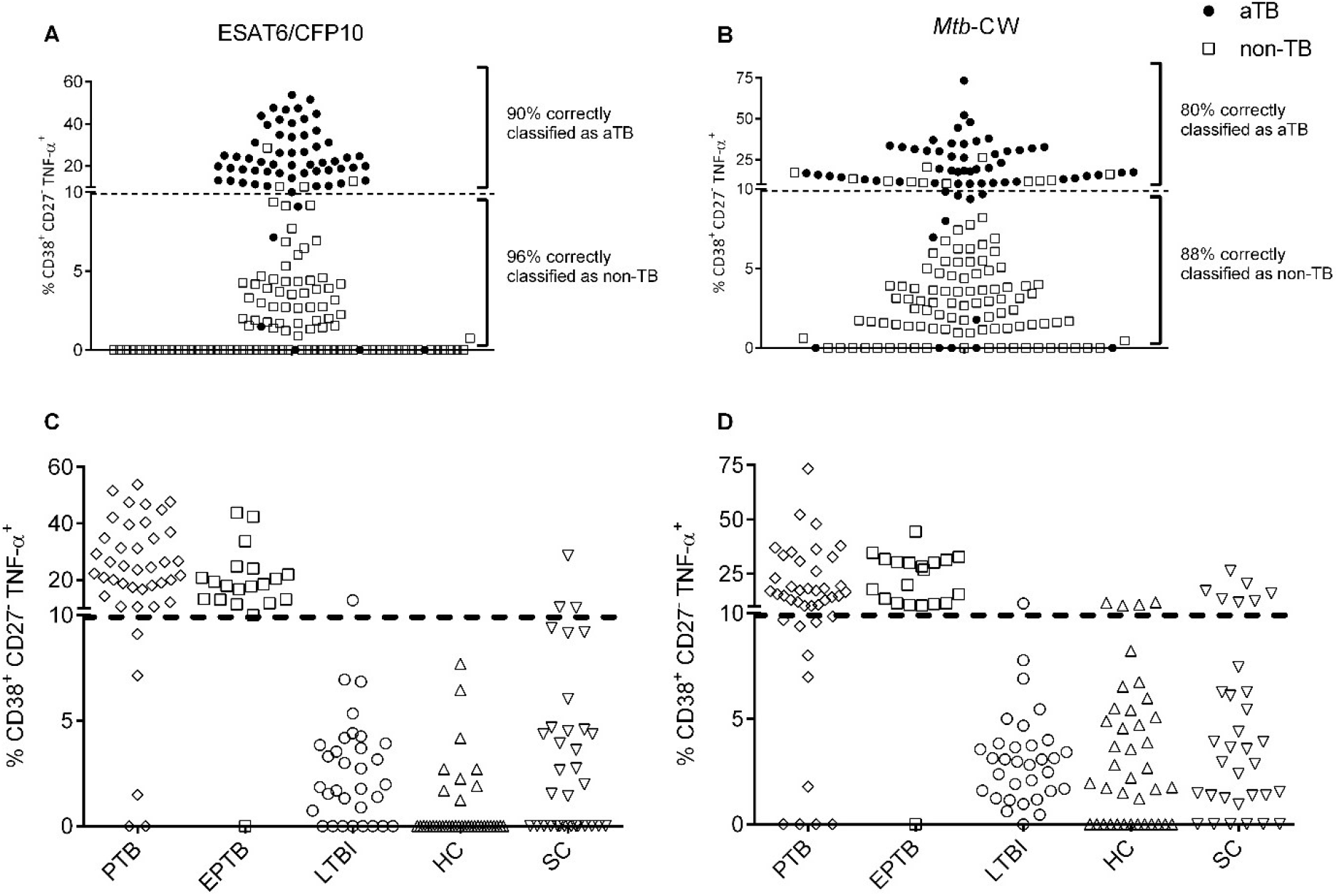
Frequencies of CD38^+^CD27^−^TNF-α^+^ in the validation cohort. Frequencies of CD38^+^CD27^−^TNF-α^+^ from 165 individuals in **(A)** ESAT6/CFP10 and **(B)** *Mtb-*CW antigen stimulated whole blood have been summarised. The predetermined cut-off value of 9.91% is indicated by the dashed line. Frequencies of CD38^+^CD27^−^TNF-α^+^ were normalised to 0% in samples with no significant CD4^+^TNF-α^+^ response. Filled circles and hollow squares correspond to individuals from aTB (PTB and EPTB) and non-TB groups (LTBI, HC, SC) respectively, after unblinding. Frequencies of CD38^+^CD27^−^TNF-α^+^ from PTB (n= 41), EPTB (n = 20), LTBI (n = 32), HC (n = 39) and SC (n = 33) upon **(C)** ESAT6/CFP10 and **(D)** *Mtb-*CW antigen have been summarized, after unblinding. Black dashed line represents the predetermined cut-off value of 9.91%.

### Identification of additional biomarkers within mono and polyfunctional CD4^+^ T cell subsets

We further aimed to evaluate the diagnostic potential of CD38^+^CD27^−^, CD38^+^ and CD27^−^ phenotypes on *Mtb*-specific CD4^+^ T cells in combination with their cytokine (IFN-ɣ, TNF-α, IL-2) secretion profiles. The analyses were blinded and only candidate biomarkers having AUC values greater than 0.875 with both ESAT6/CFP10 and *Mtb-*CW stimulation were considered. When significant responders alone were considered for analysis, all potential biomarkers had better performance with ESAT6/CFP10 than with *Mtb-*CW stimulation. The results have been summarised in Table 2 and Table E6. In agreement with earlier reports, CD38^+^IFN-ɣ^+^ correctly discriminated non-TB from aTB with sensitivity and specificity greater than 85% with both ESAT6/CFP10 and *Mtb-*CW stimulation in subjects with significant IFN-ɣ response (12, 13). CD38^+^TNF-α^+^ and CD38^+^IL-2^+^ frequencies did not have a high discriminatory power (data not shown). However, CD38^+^IFN-ɣ^+^TNF-α^+^ had marginally better AUC values when compared to CD38^+^IFN-ɣ^+^.

CD38^+^CD27^−^ phenotype on multiple additional functional outcomes had high diagnostic performance. Among all combinations, CD38^+^CD27^−^IFN-ɣ^+^TNF-α^+^ and CD38^+^CD27^−^IFN-ɣ^+^ had the best diagnostic performance when only samples with significant responses were considered. The cytokine production (after background deduction) from *Mtb*-specific CD4^+^ T cells in whole blood is shown in Figure E2. Out of the 61 aTB patients, 58 had significant CD4^+^TNF-α^+^ response, 43 had significant CD4^+^IFN-ɣ^+^ response and only 36 had significant CD4^+^IL-2^+^ response with ESAT6/CFP10 stimulation (Table 2). With *Mtb-*CW stimulation, 57, 49 and 47 had significant CD4^+^TNF-α^+^, CD4^+^IFN-ɣ^+^ and CD4^+^IL-2^+^ response, respectively (Table E6). Due to the absence of significant IFN-ɣ and IL-2 response in whole blood in a high proportion of aTB subjects, biomarkers involving IFN-ɣ or IL-2 combinations are not feasible for use in clinical settings. Thus, the robust production of TNF-α makes it the cytokine of choice for discriminating aTB from non-TB individuals.

### High prevalence of CD38^−^CD27^+^ phenotype in *Mtb*-exposed individuals without active disease

Frequencies of CD38^−^CD27^+^ phenotype on *Mtb*-specific CD4^+^ T cells defined by several functional outcomes were significantly higher in *Mtb-*exposed (LTBI, CTB and SC) subjects compared to aTB patients (Figure 4D, Figure 6 and Figure E4). Nevertheless, the considerable overlap evident between aTB and non-TB subjects precluded their use as reliable biomarkers. Since frequencies of CD38^−^CD27^+^ on *Mtb*-specific CD4^+^ T cells in CTB and SC groups were comparable with those in LTBI, these subsets could also be explored for monitoring treatment response and determining cure. Interestingly, in contrast to earlier reports (17) we did not observe CD38^−^CD27^−^ phenotype on *Mtb*-specific CD4^+^ T cells to be dominant in subjects who had completed ATT (data not shown).

**Figure 6.**
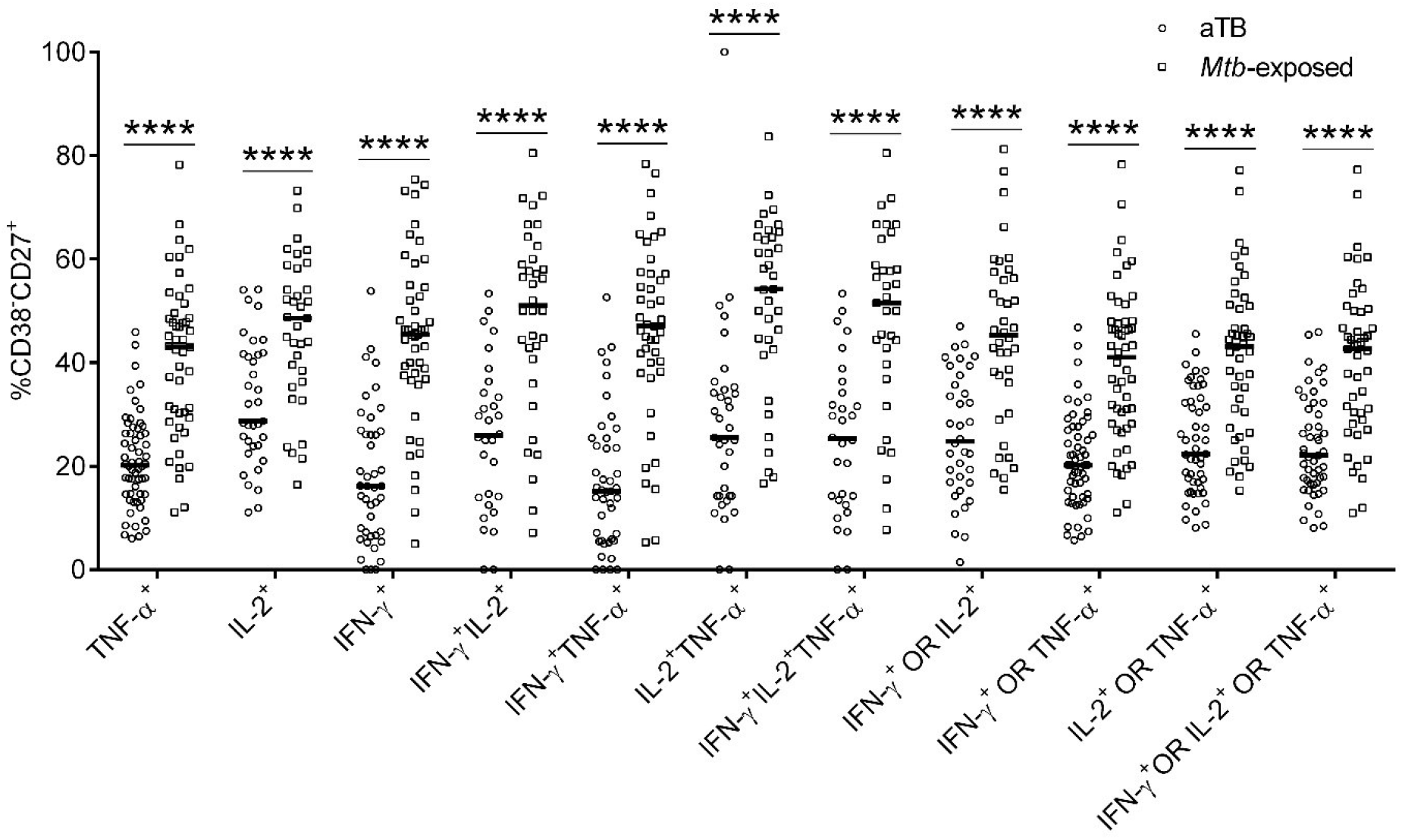
Frequencies of CD38^−^CD27^+^ on *Mtb*-specific on mono and polyfunctional CD4^+^ T cells. CD38^−^CD27^+^ frequencies on *Mtb*-specific CD4^+^ T cells upon stimulation with ESAT6/CFP10 were significantly higher in in *Mtb*-exposed individuals (LTBI and SC) compared to individuals with aTB (PTB and EPTB). Only samples with significant cytokine responses have been analysed. The number of responders for each cytokine or their combination have been summarised in Table 2 and Table E6. Mann-Whitney *U* test was used to compare the two groups for each functional outcome: ****p < 0.0001. Bars represent medians.

## DISCUSSION

In this study, we identified and validated *Mtb*-specific TNF-α secreting CD38^+^CD27^−^CD4^+^ T cells as a novel biomarker for diagnosis of aTB. We first identified this subset using PBMCs in the exploratory cohort and further validated its performance in whole blood from an independent blinded cohort. To mimic the clinical setting we classified non responders as negative for the biomarker and retained them for analyses in contrast to other biomarker studies (12, 21, 22). Our study also included control subjects with other respiratory diseases who had previously completed ATT. The biomarker displayed impressive diagnostic accuracy in correctly distinguishing aTB (both PTB and EPTB) from non-TB groups (LTBI, SC, HC and CTB), testifying to its added potential for monitoring treatment response. It was especially heartening to note the reliability of the biomarker in correctly identifying EPTB patients where the difficulty in detecting bacilli makes diagnosis particularly challenging for the clinicians. Independent studies showing high frequencies of CD38^+^ and CD27^−^ on *Mtb*-specific CD4^+^ T cells in subjects co-infected with TB and HIV, suggest the utility of this biomarker also in these individuals (13–15, 23).

CD38^+^CD27^−^CD4^+^ T cells secreting IFN-ɣ or IL-2 alone or in combination with each other or with TNF-α and CD38^+^ on polyfunctional CD4^+^ T cells had high predictive power in discriminating aTB from non-TB (Table 2 and Table E6). However, their absence in a considerable proportion of subjects precluded their choice as useful biomarkers of TB disease. Multiple previously characterised host immune biomarkers such as CD27^−^ phenotype on IFN-ɣ^+^TNF-α^+^CD4^+^ T cells (10), CD27^−^ phenotype on IFN-ɣ^+^CD4^+^ T cells (10, 24, 25), and monofunctional and polyfunctional CD4^+^ T cells (20, 26–28) did not effectively distinguish non-TB groups from aTB in our validation cohort using whole blood and had AUC values considerably below 0.875.

CD38^+^IFN-ɣ^+^, HLA-DR^+^IFN-ɣ^+^ and Ki-67^+^IFN-ɣ^+^ have shown high diagnostic performance in discriminating latent from active TB in several independent studies (12–16). CD38^+^IFN-ɣ^+^ had high predictive power when we considered only samples with significant IFN-ɣ responses (Table 2 and Table E6). Recently, Musvosvi *et al*. using whole blood showed that HLA-DR^+^ and HLA-DR^+^CD27^−^ phenotypes on antigen-specific CD4^+^ T cells secreting IFN-ɣ or TNF-α or IL-2 alone or in combination with each other had high sensitivity and specificity for TB diagnosis. Additionally they reported that a simple four colour flow cytometry panel comprising CD3, HLA-DR, IFN-ɣ and TNF-α was sufficient to discriminate latent from aTB (21). Despite their high sensitivity and specificity, the exclusion of non-responders from analyses calls into question the utility of these biomarkers in a field setting. By including all the non-responders in the analyses, our data represent a truly realistic picture of the diagnostic accuracy of the identified biomarker in a clinical setting.

Flow cytometry is today commonplace in diagnostic laboratories enabling the exploitation of host blood-based biomarkers for disease diagnosis. Our results affirm the potential of phenotypic biomarkers on *Mtb*-specific CD4^+^ T cells defined by production of TNF-α rather than IFN-ɣ or IL2 to be a superior and robust biomarker. Identification of the unique CD4^+^ T cell subset with CD38^+^CD27^−^ phenotype allowed us to exploit the potential of the abundant cytokine TNF-α to correctly identify both pulmonary and extra-pulmonary tuberculosis with high predictive values in whole blood assays that can be performed with a single laser 5-parameter flow cytometer.

## Data Availability

Study protocols, exported flow cytometry data and de-identified FCS files that underlie the results reported are available from the corresponding author upon reasonable request.

## ACKNOWLEDGMENTS

We sincerely thank all the study participants, ESIC MC & PGIMSR staff and healthcare workers of RNTCP (Revised National Tuberculosis Control Programme presently National Tuberculosis Elimination Programme (NTEP))-Hosahalli TB Unit. This work was funded by DBT-IISc Partnership Program Phase-II (BT/PR27952/INF/22/212/2018). The funding agency had no role in the study. We gratefully acknowledge Beckman Coulter India Private Limited for providing flow cytometer, antibodies, reagents and salary for MPA. We thank the Biosafety Level-3 (BSL-3) facility at IISc.

## DECLARATION OF INTERESTS

VS received grants from Department of Biotechnology, Government of India (BT/PR23695/MED/29/1195/2017 Dated 28/12/2017) and Beckman Coulter India Private Limited (SP/BECO-16-0001) outside the submitted work. MPA received salary from Beckman Coulter India Private Limited outside the submitted work.

## AUTHOR CONTRIBUTIONS

VS conceived the study, supervised and edited the manuscript. MPA designed, planned, conducted the experiments, analysed and interpreted data and wrote the manuscript. SPP assisted with PBMC processing in exploratory cohort. VSM, PC, VK, SN, SBN, N, RY were responsible for diagnosis, consenting and recruitment of patients, clinical sample collection and maintenance of case records and reports.

## ONLINE DATA SUPPLEMENT

### List of Supplementary Materials

#### Supplemental Methods

Figure E1. Gating Strategy.

Figure E2. Representative flow plots showing phenotypic characteristics of *Mtb*-specific TNF-α^+^CD4^+^ T cells upon stimulation with ESAT6/CFP10 and *Mtb*-CW antigens.

Figure E3. Analysis of cytokine secreting Mtb-specific CD4^+^ T cells.

Figure E4. Frequencies of CD38^−^CD27^+^ on *Mtb*-specific on mono and polyfunctional CD4^+^ T cells.

Table E1: Description of study participants.

Table E2: Clinical description of pulmonary tuberculosis patients included in the study.

Table E3: Clinical description of extrapulmonary tuberculosis patients included in the study.

Table E4: Clinical description of sick controls in the study.

Table E5. Summary of the antibody panels used in the study.

Table E6. Biomarker performance of CD38^+^CD27^−^ and CD38^+^ within mono- and polyfunctional CD4^+^ T cells.

## SUPPLEMENTAL METHODS

Preparation of *Mtb*-cell wall (*Mtb*-CW) antigen:

*Mtb-*CW was prepared as described previously (E1, E2). Briefly, bacteria (H37Rv) grown to late log phase (14 days) were heat killed at 80°C for 1h. Cells were pelleted at 5000 x g, washed four to five times with PBS and suspended at a concentration of 0.5g/mL in PBS containing 3mM EDTA, DNase (1mg/mL; Sigma, D5025), RNase (1mg/mL; Sigma, R4875) and protease inhibitor cocktail (Sigma, P8465). The cells were disrupted with 0.1mm zirconia beads in FastPrep-24 (MP Biomedicals, LLC) at 4.5m/s for 1min followed by 2mins rest on ice for 8 cycles. The beads were allowed to settle and the collected supernatant was sonicated in a Branson 250/450 sonifier at 30% amplitude for 2mins with 5mins rest on ice for 8 cycles. The lysate was centrifuged at 5000 x g twice to remove partially broken/unbroken cells. The supernatant was centrifuged at 27000 x g (4°C) for 1h and the cell wall pellet was resuspended and dialysed (3500 MWCO) in 10mM ammonium bicarbonate for 24h at 4°C with three buffer changes and quantified by Bradford’s assay. Protein quality of the *Mtb-*CW fraction was assessed by SDS-PAGE and silver staining.

**Figure E1.**
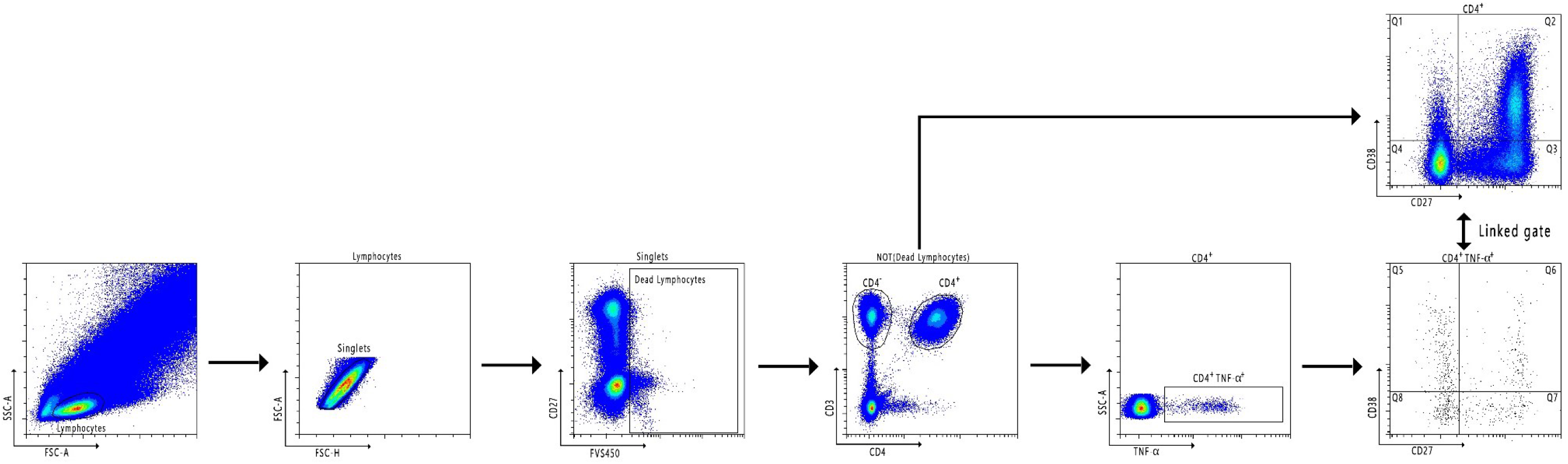
Gating Strategy. Lymphocytes were obtained using Forward Scatter Area (FSC-A) versus Side Scatter Area (SSC-A) followed by singlets from Forward Scatter-Height and FSC-A. Dead lymphocytes were excluded and the CD3^+^CD4^+^ population was selected from live lymphocytes that was obtained from the NOT (Dead Lymphocytes) population. TNF-α producing CD4^+^ T cells were selected and were displayed on CD27 vs CD38. The same quadrant gate on CD27 vs CD38 on total CD4^+^ T cells was applied to CD27 vs CD38 on CD4^+^TNF-α^+^ T cells using the “Link Gate” feature in CytExpert v2.0. CD38^+^TNF-α^+^: (Q5 + Q6), CD27^−^TNF-α^+^: (Q5+Q7), CD38^+^CD27^−^TNF-α^+^: Q5 and CD38^−^CD27^+^TNF-α^+^: Q7.

**Figure E2.**
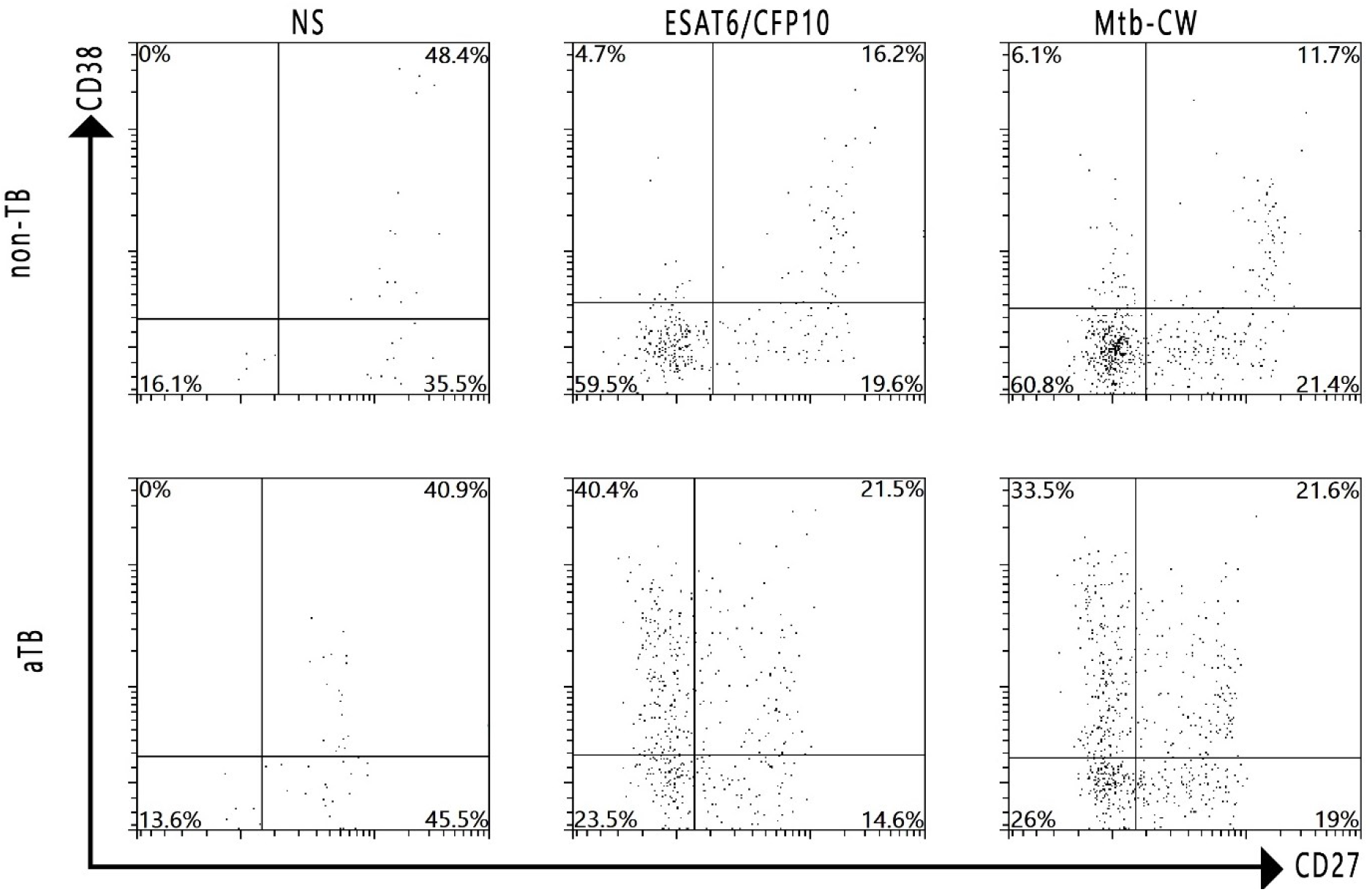
Representative flow plots showing phenotypic characteristics of *Mtb*-specific TNF-α^+^CD4^+^ T cells upon stimulation with ESAT6/CFP10 and *Mtb*-CW antigens.

**Figure E3.**
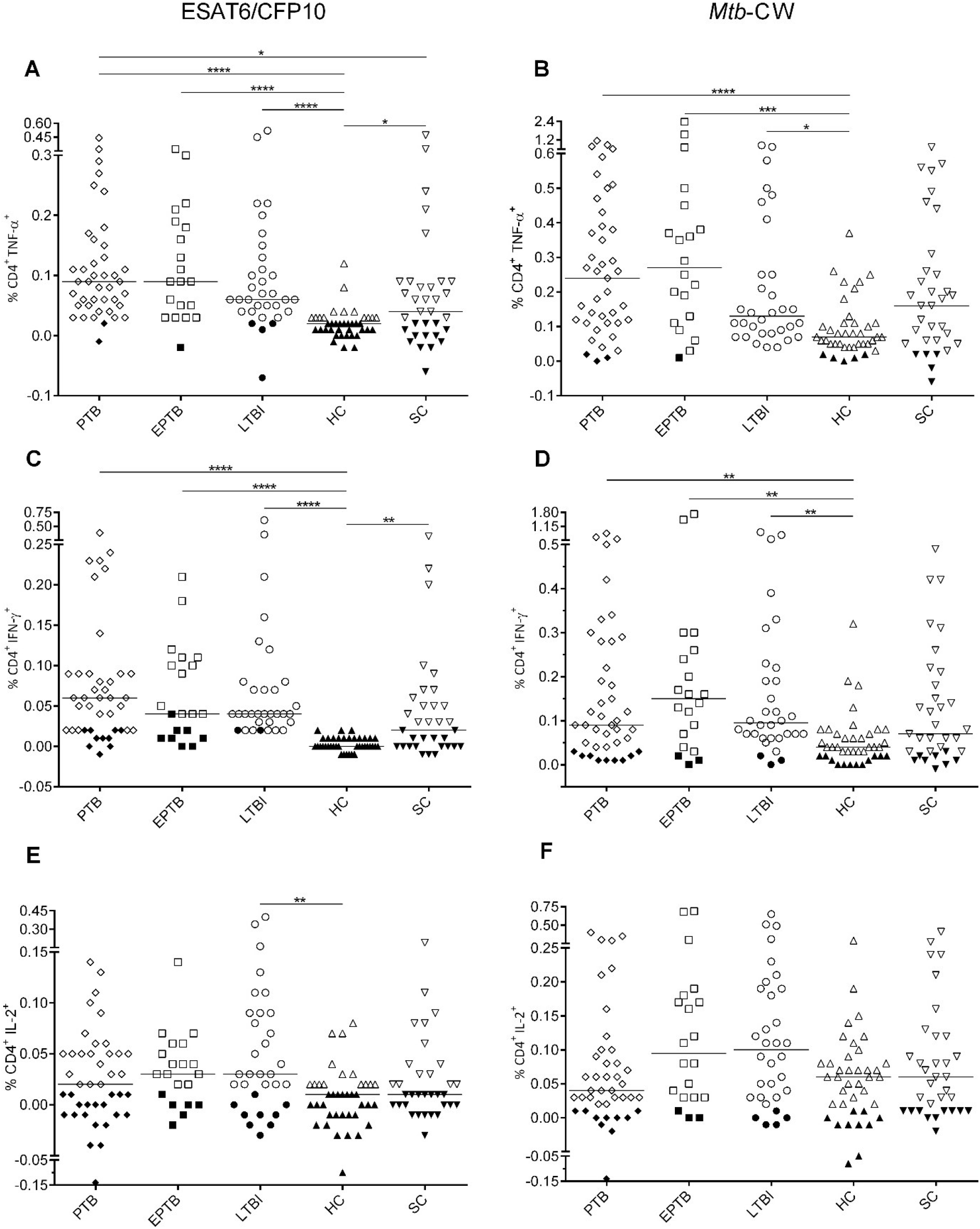
Analysis of cytokine secreting Mtb-specific CD4^+^ T cells. Whole blood from PTB (n= 41), EPTB (n = 20), LTBI (n = 32), HC (n = 39) and SC (n = 33) were stimulated with ESAT6/CFP10 **(A, C, E)** or *Mtb*-CW antigen **(B, D, F)**. Frequencies of CD4^+^TNF-α^+^ **(A, B)**, CD4^+^IFN-ɣ^+^ **(C, D)**, CD4^+^IL-2^+^ **(E, F)** are reported after subtracting the unstimulated response from the stimulated tubes. Kruskal-Wallis test followed by Dunn’s post-test was used to compare the groups: *p < 0·05; **p < 0·01; ***p < 0·001; ****p < 0·0001. Samples with no-significant cytokine response are highlighted using filled symbols. Bars represent medians.

**Figure E4.**
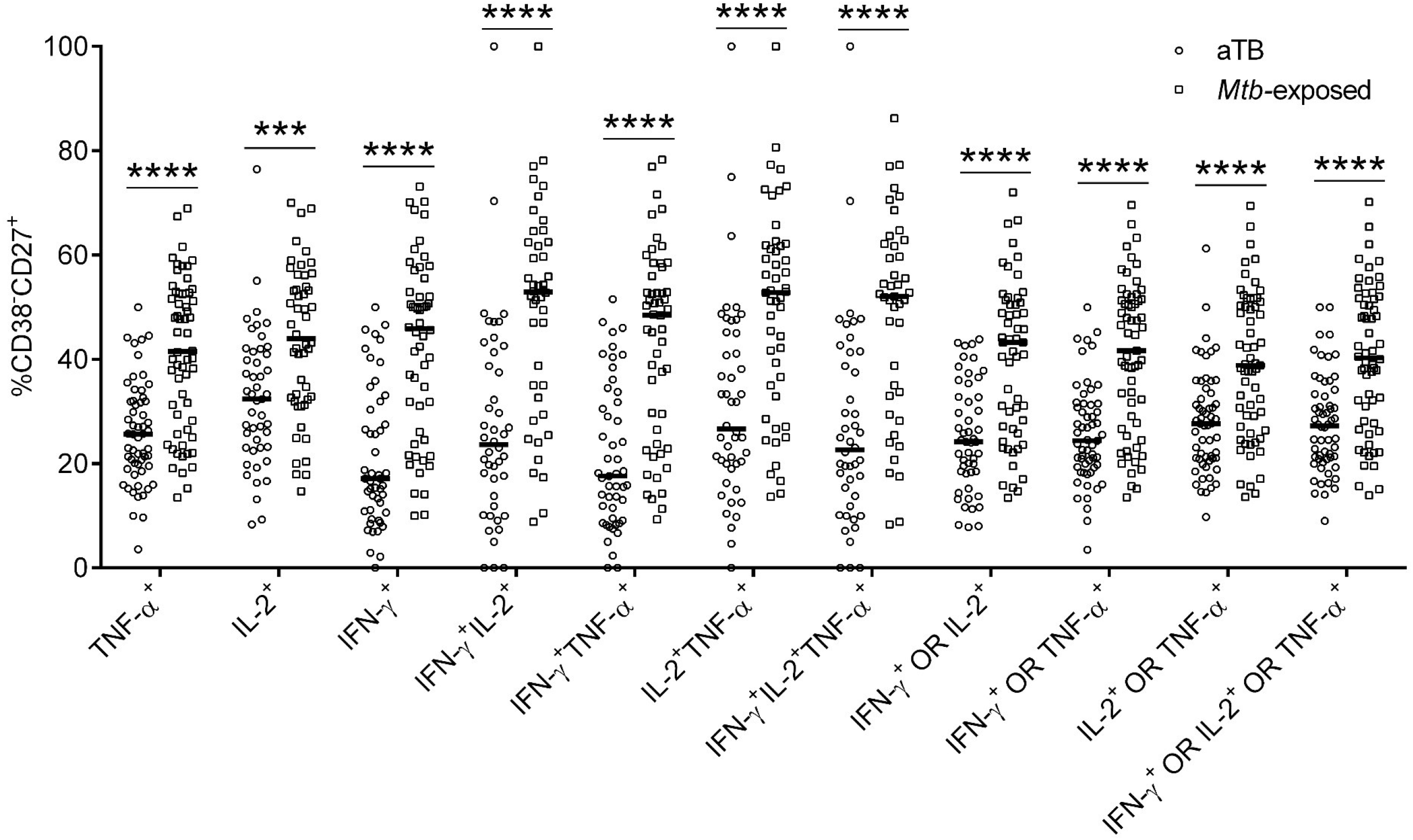
Frequencies of CD38^−^CD27^+^ on *Mtb*-specific on mono and polyfunctional CD4^+^ T cells. CD38^−^CD27^+^ frequencies on *Mtb*-specific CD4^+^ T cells upon stimulation with *Mtb*-CW were significantly higher in in *Mtb*-exposed individuals (squares) (LTBI and SC) compared to individuals with aTB (circles) (PTB and EPTB). Only samples with significant cytokine responses have been analysed. The number of responders for each cytokine or their combination have been summarised in Table 2 and Table E6. Mann-Whitney *U* test was used to compare the two groups for each functional outcome: ***p < 0.001, ****p < 0.0001. Bars represent medians.

**Table E1:** Description of study participants.

|  |  |
| --- | --- |
| Exploratory cohort (EC) | 79 participants recruited were categorised into four groups: 15 having PTB (2 excluded from analysis – 1 HBV+, 1 insufficient blood volume), 15 with EPTB (2 excluded from analysis – insufficient blood volume) , 16 cured patients (1 excluded from analysis – TB relapse) and 33 healthy individuals out of which 16 were found to be latently infected with *Mtb* (1 excluded from analysis – insufficient blood volume). |
| Validation cohort (VC) | 166 participants recruited randomly and were categorised post analysis as follows: 41 having PTB, 21 with EPTB (1 excluded from analysis – HIV+), 33 sick controls previously treated for TB and 71 healthy individuals out of which 32 were found to be latently infected with *Mtb*. |
| Pulmonary tuberculosis patients (PTB) | Subjects recruited were either positive for sputum AFB or for PCR (Xpert-MTB/RIF - Cepheid) or both. Subsequent culture of sputum revealed all recruited cases to be culture positive. In most cases, First Line – Line Probe Assay (GenoType MTBDRplus V2 - Hain Lifescience GmbH) was performed to detect Rifampicin and Isoniazid resistance. |
| Extrapulmonary tuberculosis patients (EPTB) | EPTB cases were confirmed by at least one of the following tests in addition to the clinical diagnosis: ZN stain (aspirates/biopsies), FNAC/FNAB (aspirates/ biopsies), PCR - Xpert-MTB/RIF (body fluids/ aspirates/ biopsies), or predominant lymphocyte infiltration and ADA levels > 40U/mL (pleural fluid and ascites). Disease sites included lymph node, pleural fluid, brain, urinary bladder/ureter, ovary/fallopian tube, |
|  | peritoneal cavity, intestine and eye. Extrapulmonary infections associated with pulmonary infections were not recruited. |
| Latently infected subjects (LTBI) and healthy controls (HC) | Health care workers at ESIC MC & PGIMSR, Revised National Tuberculosis Control Programme (RNTCP) presently National Tuberculosis Elimination Programme (NTEP) staff of Hosahalli TB unit and *Mtb* exposed household contacts with no symptoms of the disease and no previous history of tuberculosis were screened for latent TB infection. IGRA+ subjects or individuals with significant ESAT6/CFP10-specific IFN-γ<sup>+</sup>CD4<sup>+</sup> T cell responses in 20hr stimulated whole blood were classified as latent TB. The non-responders were categorised as *healthy controls (HC)* and in the exploratory cohort were not processed further. |
| Cured TB subjects (CTB) | Recently treated PTB patients who had completed ATT in the past 1 – 2 months with sputum negative for smear AFB and culture were recruited. |
| Previously treated sick controls (SC) | Patients who had previously completed ATT for PTB at least 1 year prior and presenting themselves at the hospital with chest/lung infection having clinical symptoms of TB were recruited. They were all sputum smear negative and completely recovered from their symptoms within 10 - 15 days of antibiotic/antifungal/antiviral treatment. All these sputum cultures were negative. |

**Table E2:** Clinical description of pulmonary tuberculosis patients included in the study.

| ID | Age | Sex | Symptoms | AFB | PCR (Rif) | FL-LPA (Rif, Inh) | Culture |
| --- | --- | --- | --- | --- | --- | --- | --- |
| **Exploratory cohort** |  |  |  |  |  |  |  |
| P1 | 65 | M | cough, weight loss, fatigue, extensive non homogenous opacities in right lung | 1+ | + (S) | .. | + |
| P2 | 35 | M | cough, fever, weight loss, night sweats, nodules in the lungs | 1+ | + (S) | + (S, S) | + |
| P3 | 29 | F | cough, fever, weight loss, lassitude, chest pain, left pleural effusion | 3+ | + (S) | + (S, S) | + |
| P4 | 49 | M | cough, fever, chest pain, left pleural effusion | 3+ | + (S) | + (S, S) | + |
| P5 | 50 | M | cough, fever, weight loss, night sweats, chills, consistent non homogenous opacities in the lungs | 2+ | + (S) | .. | + |
| P6 | 44 | M | cough, fever, weight loss, hemoptysis | 2+ | + (S) | .. | + |
| P7 | 55 | F | cough, fever, weight loss, lassitude, throat pain, cavitory lesion in the left lower lobe | 2+ | + (S) | + (S, S) | + |
| P8 | 30 | F | cough, fever, weight loss, weakness, fatigue, nodular densities in the left lung | 2+ | + (S) | + (S, S) | + |
| P9 | 44 | F | cough, fever, weight loss, lassitude, chest pain, cavitation in right middle lobe | 3+ | + (S) | + (S, S) | + |
| P11 | 65 | M | cough, fever, weight loss, lassitude, bilateral cavitory lesions in the lower lobes | 2+ | + (S) | + (S, S) | + |
| P12 | 48 | M | cough, fever, chest pain, hemoptysis, pleural effusion in left lung, multiple cavitory lesions in right upper and middle lobes | 3+ | .. | .. | + |
| P13 | 28 | M | cough, weight loss, chest pain, fatigue, cavitory lesion in right upper lobe | 2+ | + (S) | + (S, S) | + |
| P14 | 17 | F | productive cough, evening rise of temperature, cavitation in right lower and middle lobes | 2+ | + (R) | *SL LPA:*<br>+ (FQ: S, K: R) | + |
| **Validation cohort** |  |  |  |  |  |  |  |
| P15 | 48 | M | intermittent fever, cough, chest pain, weight loss, loss of appetite, bilateral cavitation in lower lobes | 3+ | + (S) | + (S, S) | + |
| P16 | 48 | M | cough, fever, weight loss, night sweats, loss of appetite, weakness, consistent non homogenous opacities in the lungs, right pleural effusion | 3+ | + (S) | + (S, S) | + |
| P17 | 45 | M | cough, fever with chills, weight loss, headache, nausea, loss of appetite, nodular densities in both the lungs | 3+ | + (S) | + (S, S) | + |
| P18 | 41 | M | cough, fever, hemoptysis, chest pain, weight loss, loss of appetite, nodular densities in both the lungs, multiple cavitory lesions in right lung | 3+ | + (S) | .. | + |
| P19 | 28 | F | persistent cough, pain in the right eye (chronic uveitis), consistent non-homogenous opacities the lungs, nodules in the right upper lobe | Neg | + (S) | .. | + |
| P20 | 52 | M | cough, fever, weight loss, chest pain, bilateral cavitation in middle lobes | 3+ | + (S) | + (S, S) | + |
| P21 | 42 | M | cough, fever, weight loss, night sweats, cavitation in right middle and lower lobes, left pleural effusion | 1+ | + (S) | + (S, S) | + |
| P22 | 40 | F | cough, fever, weight loss consistent non homogenous opacities in the lung, pleural effusion | 1+ | + (S) | + (S, S) | + |
| P23 | 41 | M | evening rise of temperature, chest pain, weight loss, loss of appetite, night sweats | 2+ | + (S) | + (S, S) | + |
| P24 | 63 | M | cough, fever, weight loss, loss of appetite, cavitory lesion in the left middle lobe | 3+ | + (S) | .. | + |
| P25 | 50 | M | cough, fever, hemoptysis, weight loss, bilateral cavitation in middle lobes | 3+ | + (S) | + (S, S) | + |
| P26 | 52 | M | cough, fever with chills, weight loss, chest pain, loss of appetite, complete destruction of left lung, non-homogenous opacities in the right lung | 3+ | + (S) | + (S, S) | + |
| P27 | 37 | M | cough, fever, weight loss, loss of appetite, small nodular densities in the lungs | 1+ | + (S) | + (S, S) | + |
| P28 | 22 | M | productive cough, fever | 3+ | + (S) | + (S, S) | + |
| P29 | 58 | M | cough, fever, weight loss, chest pain, cavitation in left middle and upper lobes | 2+ | + (S) | + (S, S) | + |
| P30 | 60 | M | cough, fever, weight loss, chest pain, loss of appetite, calcified nodules and cavitory lesions in the left lower lobe | 3+ | + (S) | + (S, S) | + |
| P31 | 39 | M | cough, fever, cavitory lesions in the left upper and middle lobe | 1+ | + (S) | + (S, S) | + |
| P32 | 45 | M | cough, fever, weight loss, headache, body pain, lassitude | Neg | + (S) | + (S,R) | + |
| P33 | 46 | F | productive cough, chest pain, left pleural effusion | Neg | + (S) | + (S,R) | + |
| P34 | 26 | F | cough, chest pain, weight loss, loss of appetite, headache, lassitude, cavitation in upper right lobe | 3+ | + (S) | + (S, S) | + |
| P35 | 50 | M | persistent cough, weight loss, multiple cavitory lesions in the left lung | 2+ | + (S) | + (S, S) | + |
| P36 | 26 | F | cough, fever, weight loss, loss of appetite, consistent non homogenous opacities in the lung | 3+ | + (S) | + (S, S) | + |
| P37 | 44 | F | cough, fever accompanied with chills, night sweat, loss of appetite, calcified nodules in the left lower and middle lobe | 2+ | + (S) | + (S, S) | + |
| P38 | 49 | M | cough, weight loss, lassitude, pulmonary infiltrates in the right lung | 3+ | + (S) | + (S, S) | + |
| P39 | 30 | F | fever, chills, night sweats, weight loss, loss of appetite, chest pain, multiple cavitory lesions in the left middle and lower lobes | 3+ | + (S) | + (S, S) | + |
| P40 | 48 | M | cough, weight loss, loss of appetite, lassitude, lung apices have diffuse fibro cavitory changes and areas of reticular opacities on CT scan | 3+ | + (S) | + (S, S) | + |
| P41 | 55 | M | cough, evening rise of temperature, weight loss, chills, fatigue, complete destruction of left lung, non-homogenous opacities in the right lung | 3+ | + (S) | + (S, S) | + |
| P42 | 44 | M | persistent cough, severe weight loss, night sweats<br>cavitation in right upper lobe, nodular densities in right middle and lower lobes | 1+ | + (S) | + (S, S) | + |
| P43 | 60 | F | cough, fever, weight loss, loss of appetite, lassitude, multiple nodular densities in the lung, left pleural effusion | 3+ | + (S) | + (S,R) | + |
| P44 | 39 | M | cough, evening rise of temperature, weight loss, loss of appetite, hemoptysis, lassitude, non-homogenous opacities in the lung | 2+ | + (S) | + (S, S) | + |
| P45 | 27 | M | cough, weight loss, loss of appetite, lassitude, multiple cavitory lesions in the right lung | 3+ | + (S) | + (S, S) | + |
| P46 | 34 | M | cough, intermittent fever, weight loss, loss of appetite, body pain, night sweats, giddiness, lassitude, nodular densities in both the lungs | 3+ | + (S) | + (S, S) | + |
| P47 | 32 | F | cough, fever, weight loss, loss of appetite, night sweats, left pleural effusion | 3+ | + (S) | + (S, S) | + |
| P48 | 50 | M | cough, fever, chills, chest pain, joint pain,<br>loss of appetite, cavitation in right lower<br>lobe | 3+ | + (S) | + (S, S) | + |
| P49 | 39 | M | cough, fever, weight loss, chest pain, night<br>sweats, bilateral cavitation in upper lobes,<br>right pleural effusion | 3+ | + (S) | + (S, S) | + |
| P50 | 50 | M | cough, fever, weight loss, loss of appetite,<br>lassitude, complete destruction of left lung,<br>left pleural effusion | 1+ | + (S) | + (S, S) | + |
| P51 | 56 | M | cough, evening rise of temperature, weight<br>loss, night sweats, fatigue, body pain,<br>bilateral cavitory lesion in upper lobes | 3+ | + (S) | + (S, S) | + |
| P52 | 41 | M | cough, evening rise of temperature, weight<br>loss, loss of appetite, hemoptysis, nodular<br>densities in left middle and lower lobes,<br>complete destruction of right lung | 2+ | + (S) | + (S, S) | + |
| P53 | 31 | F | cough, fever, weight loss, loss of appetite,<br>lassitude | 2+ | + (S) | + (S, S) | + |
| P54 | 49 | F | cough, fever, weight loss, loss of appetite,<br>multiple cavitory lesions in the left lung | 3+ | + (S) | + (S, S) | + |
| P55 | 50 | M | cough, fever, chest pain, night sweats,<br>cavitation in left upper lobe | 3+ | + (S) | + (S, S) | + |
| P56 | 48 | M | cough, intermittent fever, weight loss, night sweats, lassitude, chest pain, multiple cavitory lesions in the left lung | 2+ | + (S) | + (S, S) | + |

**FL-LPA** First line - Line probe assay **Rif** Rifampicin susceptibility
**SL-LPA** First line - Line probe assay **Inh** Isoniazid susceptibility
**CT scan** Computed tomography scan **FQ** Fluoroquinolones susceptibility
**AFB** acid fast bacilli **K** Kanamycin susceptibility
.. Test not done/Data not available + Positive — Negative
(S) Sensitive (R) Resistant

**Table E3:** Clinical description of extrapulmonary tuberculosis patients included in the study.

| ID | Age | Sex | Symptoms | Tests/Result | Final Diagnosis |
| --- | --- | --- | --- | --- | --- |
| **Exploratory cohort** |  |  |  |  |  |
| EP1 | 33 | M | swelling in the right posterior part of the neck | *FNAC* (right posterior cervical lymph node aspirate): Cytological features are suggestive of granulomatous lymphadenitis of tuberculous etiology. *ZN stain for AFB*: positive. | Lymph Node Tuberculosis |
| EP2 | 34 | M | intermittent fever, burning micturition, abdominal pain, weight loss | *CT scan* (KUB region): Left sided polynephritis and ureteritis with multiple left ureteric strictures and moderate left hydronephrosis. Right lower ureter wall thickening suggesting of ureteritis with no ureteric narrowing or hydronephrosis. Small capacity urinary bladder with irregular thickening of bladder wall. Multiple enlarged para-aortic lymph node. Pleural thickening/effusion on right side. *Xpert-MTB/RIF* (urine): MTB detected; Rif sensitivity: indeterminate. | Tuberculous ureteritis |
| EP5 | 44 | M | headache,<br>intermittent fever,<br>weight loss | MRI (brain): 4x4mm small ring enhancing lesion surrounding parenchymal edema on parietal corona radiata region, probably of Koch's. *Xpert-MTB/RIF* (CSF) - MTB detected; Rif sensitive. | TB meningitis |
| EP6 | 37 | M | fever, weight loss,<br>lassitude,<br>swelling, pain and<br>tenderness in left<br>side of the neck | *FNAC* (left cervical lymph node aspirate): Cytological features are suggestive of reactive lymphadenitis of tuberculous etiology. *ZN stain for AFB*: negative.<br>*FNAB* (left cervical lymph node biopsy): Histopathological features are suggestive of caseating granulomatous lymphadenitis of tuberculous etiology. *ZN stain for AFB*: positive. | Lymph Node<br>Tuberculosis |
| EP7 | 64 | M | evening rise of<br>temperature,<br>lassitude, loss of<br>appetite, weight<br>loss, pleural effusion in right lung | Seronegative.<br>*Biochemistry* (pleural fluid): ADA - 91U/ml.<br>*Cytology* (pleural fluid): 98% lymphocytes. | Tuberculous pleural<br>effusion |
| EP8 | 21 | F | swelling and pain in the right anterior side of the neck | *FNAC* (right anterior cervical lymph node aspirate): Cytological features are suggestive of granulomatous lymphadenitis of Koch's etiology. *ZN stain for AFB*: positive.<br>*FNAB* (right anterior cervical lymph node biopsy): Histopathological features are suggestive of caseating granulomatous lymphadenitis of Koch's etiology. *ZN stain for AFB*: positive. | Lymph Node Tuberculosis |
| EP9 | 35 | M | evening rise of temperature, lassitude, weight loss, pleural effusion in left lung | *Sputum AFB*: negative.<br>*Xpert-MTB/RIF* (pleural fluid): MTB detected; Rif sensitive. | Tuberculous pleural effusion |
| EP10 | 22 | F | fever, abdominal pain, loss of appetite, irregular menstrual cycle | *USG* (abdomen): Cystic mass in the left adnexa. *FNAB* (tubo-ovarian mass): Histopathological features suggestive of caseating granulomatous lesion of tuberculous etiology. *ZN stain for AFB*: positive. | Ovarian Tuberculosis |
| EP11 | 31 | F | swelling in right anterior side of the neck | *FNAC* (right anterior cervical lymph node aspirate): Cytological features are suggestive of caseating necrotising granulomatous lymphadenitis of Koch's etiology. *ZN stain for AFB*: negative. | Lymph Node Tuberculosis |
| EP12 | 37 | M | abdominal pain, loose stools, vomiting, weight loss, evening rise of temperature, lassitude, giddiness | *CECT* (abdomen) - Diffuse wall thickening at terminal ileum, ileocecal junction, caecum and ascending colon. Adenopathy in adjacent mesenteric lymph nodes. Features suggestive of tuberculosis. *Chest Xray*: normal. *Xpert-MTB/RIF* (ileocecal mass): MTB detected; Rif sensitive. | Ileocecal tuberculosis |
| EP13 | 57 | F | fever, headache,<br>blurred vision,<br>chronic uveitis | *IGRA*: positive. *Xpert-MTB/RIF* (aqueous humour): MTB detected; Rif sensitive. | Ocular tuberculosis |
| EP14 | 45 | F | swelling in<br>midline neck<br>region - anterior<br>part | *CECT* (neck) - Multiple inflamed lymph nodes in the media sternum region<br>*FNAC* (pus from lymph nodes): Cytological features suggestive of necrotising lesion of tuberculous etiology. *ZN stain for AFB*: negative.<br>*Xpert-MTB/RIF* (pus from anterior neck region): MTB detected; Rif sensitive. | Lymph Node<br>Tuberculosis |
| EP15 | 38 | F | cough, fever,<br>weight loss,<br>swelling and pain<br>at the base of the<br>neck | *Chest Xray*: normal. *Xpert-MTB/RIF* (sputum): MTB not detected.<br>*FNAC* (left supraclavicular lymph node aspirate): Cytological features are suggestive of granulomatous lymphadenitis of tuberculous etiology.<br>*ZN stain for AFB*: positive. | Lymph Node<br>Tuberculosis |

| Validation cohort |  |  |  |  |  |
| --- | --- | --- | --- | --- | --- |
| EP16 | 44 | F | mild fever, night sweats, swelling in right anterior part of the neck | *FNAC* (right anterior cervical lymph node aspirate): Cytological features are suggestive of caseating granulomatous lymphadenitis of Koch's etiology. *ZN stain for AFB*: positive. | Lymph node tuberculosis |
| EP17 | 48 | F | loss of appetite, night sweats, throat pain, cough, swelling in the right posterior part of the neck | *FNAC* (right posterior cervical lymph node aspirate): Cytological features are suggestive of granulomatous lymphadenitis of Koch's etiology. *ZN stain for AFB*: negative.<br>*FNAB* (right posterior cervical lymph node biopsy): Histopathological features suggestive of granulomatous lymphadenitis of Koch's etiology. *ZN stain for AFB*: negative. | Lymph node tuberculosis |
| EP18 | 29 | F | evening rise of temperature, night sweats, weight | *FNAC* (right posterior cervical lymph node aspirate): Cytological features are suggestive of caseating granulomatous lymphadenitis of | Lymph node tuberculosis |
|  |  |  | loss, swelling in the right posterior part of the neck | tuberculous etiology. *ZN stain for AFB*: negative.<br>*Xpert-MTB/RIF* (lymph node aspirate): MTB detected; Rif sensitive. |  |
| EP19 | 36 | M | pain and swelling in the right side of the neck | *FNAC* (right cervical lymph node aspirate): Cytological features are suggestive of caseating granulomatous lymphadenitis of Koch's etiology.<br>*ZN stain for AFB*: negative. | Lymph node tuberculosis |
| EP20 | 41 | M | cough, fever, headache, difficulty in breathing, abdominal pain, vomiting | *Sputum AFB*: negative.<br>*Cytology* (pleural fluid): 98% lymphocytes. *Xpert-MTB/RIF* (pleural fluid): MTB detected; Rif sensitive. | Tuberculous Pleural Effusion |
| EP21 | 27 | F | fever, weight loss, loss of appetite, | *Chest Xray*: left pleural effusion. *Xpert-MTB/RIF* (pleural fluid): MTB not detected. | Lymph node tuberculosis |
|  |  |  | swelling, pain and tenderness in the left posterior part of the neck | *FNAC* (left cervical posterior lymph node aspirate): Cytological features are suggestive of caseating granulomatous lymphadenitis of Koch's etiology. *ZN stain for AFB*: positive. |  |
| EP22 | 38 | M | swelling, pain and tenderness in the midline neck region | *FNAC* (lymph node aspirate): Cytological features are suggestive of caseating tuberculous lesion. *ZN stain for AFB*: positive. | Lymph node tuberculosis |
| EP23 | 47 | M | cough, fever, difficulty in breathing | Seronegative. *Chest Xray*: right pleural effusion.<br>*Biochemistry* (pleural fluid): ADA - 91U/ml. *Cytology* (pleural fluid): 98% lymphocytes. | Tuberculous Pleural Effusion |
| EP24 | 32 | F | pain and swelling in the left anterior side of the neck | *FNAC* (left cervical lymph node aspirate): Cytological features are suggestive of caseating granulomatous lesion of tuberculous etiology.<br>*ZN stain for AFB*: positive. | Lymph node tuberculosis |
| EP25 | 30 | F | fever, chills,<br>abdominal pain,<br>weight loss,<br>weakness | *CT scan* (abdomen): Moderate ascites in peritoneal cavity possibly due to Koch's.<br>*Biochemistry* (ascites fluid): ADA - 64·3U/ml. *Cytology* (ascites fluid): 98% lymphocytes.<br>*Xpert-MTB/RIF* (ascites fluid): MTB detected; Rif sensitive. | Abdominal tuberculosis |
| EP26 | 20 | F | pain and swelling<br>in the right side of<br>the neck | *FNAC* (right axillary lymph nodes aspirate): Cytological features are suggestive of caseating tuberculous lymphadenitis. *ZN stain for AFB*: positive. | Lymph node tuberculosis |
| EP27 | 30 | M | pain and swelling<br>in the right side of<br>the jaw | *FNAC* (submandibular lymph node aspirate): Cytological features are suggestive of granulomatous lymphadenitis of Koch's etiology. *ZN stain for AFB*: negative. *Xpert-MTB/RIF* (lymph node aspirate): MTB not detected. | Lymph node tuberculosis |
| EP28 | 21 | M | severe pain,<br>tenderness, | *USG* (Neck): Significant Lymphadenopathy Present In The Left Side Of The Neck. | Lymph node tuberculosis |
|  |  |  | swelling in the left side of the neck | *FNAC* (Left Cervical Lymph Node): Cytological Features Are Suggestive Of Caseating Tuberculous Lymphadenitis. *ZN Stain For AFB*: Positive. |  |
| EP29 | 27 | F | swelling and pain in the right supraclavicular region, evening rise of temperature, headache, lassitude | *USG* (neck): Matted lymphadenopathy (supraclavicular lymph nodes) of granulomatous etiology.<br>*FNAC* (right supraclavicular lymph node): cytological features are suggestive of granulomatous lesion of tuberculous etiology. *ZN stain for AFB*: positive | Lymph node tuberculosis |
| EP30 | 34 | F | cough, fever, loss of appetite, vomiting, lassitude, swelling | *FNAC* (supraclavicular lymph node aspirate): cytological features are suggestive of granulomatous lymphadenitis of Koch's etiology. *ZN stain for AFB*: negative | Lymph node tuberculosis |
|  |  |  | in the right supraclavicular region | *Xpert-MTB/RIF* (supraclavicular lymph node aspirate): MTB detected; Rif sensitive |  |
| EP31 | 50 | M | cough, fever, weight loss, pain in lower abdomen, burning micturition | *Chest Xray*: normal. *Sputum AFB*: negative.<br>*CT scan* (KUB): Thickened ureteral wall and calyx with no evidence of hydronephrosis, decreased bladder capacity. *Xpert-MTB/RIF* (urine): MTB detected; Rif sensitive. | Bladder tuberculosis |
| EP32 | 25 | F | pain and swelling in the left anterior side of the neck, weight loss | *USG* (neck): Multiple inflamed lymph nodes favouring tuberculosis<br>*FNAC* (left cervical lymph node): Cytological features are suggestive of granulomatous lymphadenitis of Koch's etiology. *ZN stain for AFB*: negative. | Lymph node tuberculosis |
| EP33 | 49 | M | fever accompanied with | *Chest Xray*: left pleural effusion. *Xpert-MTB/RIF* (pleural fluid): MTB not detected. | Tuberculous Pleural Effusion |
|  |  |  | chills late in the evening, weight loss, night sweats, chest pain, difficulty in breathing | *Biochemistry* (pleural fluid): ADA - 85U/ml. *Cytology* (pleural fluid): 98% lymphocytes. |  |
| EP34 | 18 | F | evening rise of temperature, swelling and pain in the right anterior part of the neck | *FNAC* (right cervical lymph node): Cytological features are suggestive of granulomatous lymphadenitis of Koch's etiology. *ZN stain for AFB*: negative. | Lymph node tuberculosis |
| EP35 | 31 | M | severe pain, tenderness, | *CT scan* (chest): Multiple inflamed lymph nodes on the left anterior chest wall favouring tuberculosis. | Lymph node tuberculosis |
|  |  |  | swelling in the left<br>anterior chest wall | *FNAC* (right humeral lymph node): Cytological features are suggestive of granulomatous lesion of Koch's etiology. *ZN stain for AFB*: positive. |  |

**CECT scan** Contrast Enhanced CT scan
**FNAC** Fine needle aspirate cytology
**MRI** Magnetic Resonance Imaging
**FNAB** Fine needle aspirate biopsy
**USG** Ultrasound scan
**ADA** Adenosine deaminase
**IGRA** Interferon- $\gamma$ release assay
**ZN** Ziehl-Neelson

**Table E4:** Clinical description of sick controls in the study.

| ID | Age | Sex | Symptoms | AFB | PCR | Culture |
| --- | --- | --- | --- | --- | --- | --- |
| SC1 | 38 | M | fever, chills, cough, night sweats, vomiting, non-homogenous opacities in the lungs | — | .. | — |
| SC2 | 32 | F | fever, cough, night sweats, malaise, lassitude, pain in the left side of the chest, nodular granules in the right lung, pleural effusion in the left lung | — | .. | — |
| SC3 | 45 | M | productive cough, fever, Xray suggestive of acute bronchitis | — | .. | — |
| SC4 | 40 | F | persistent cough, fever, loss of appetite, body pain, headache, pulmonary infiltrates in the left lung | — | .. | — |
| SC5 | 39 | M | cough, fever, chest pain, right pleural effusion | — | .. | — |
| SC6 | 47 | M | cough, evening rise of temperature, loss of appetite | — | .. | — |
| SC7 | 44 | F | cough, intermittent fever, breathlessness, difficulty in breathing, complete destruction of right lung | — | .. | — |
| SC8 | 50 | M | persistent cough, fever, malaise, lassitude, cavitation in left upper lobe | — | .. | — |
| SC9 | 49 | F | cough, fever, weight loss, lassitude | — | .. | — |
| SC10 | 44 | M | cough, fever, nodules in the right lung | — | .. | — |
| SC11 | 51 | M | productive cough, chest pain, breathlessness, Xray suggestive of bronchial asthma and COPD | — | .. | — |
| SC12 | 43 | F | cough, fever, congestion, loss of appetite | — | .. | — |
| SC13 | 47 | M | cough, fever, chest pain, weight loss, night sweats, weakness, left pleural effusion, cavitory lesions and nodules in the right lung | — | — | — |
| SC14 | 51 | M | cough, chest pain, Xray suggestive of acute infective exacerbation of COPD | — | — | — |
| SC15 | 23 | F | cough, fever, night sweats, left pleural effusion | — | .. | — |
| SC16 | 37 | F | cough, fever, loss of appetite | — | .. | — |
| SC17 | 39 | M | cough, fever, chest pain, bilateral cavitory lesions in upper and middle lobes | — | — | — |
| SC18 | 36 | M | cough, fever, weight loss, congestion, breathing difficulty, consistent non-homogenous opacities in the lungs | — | .. | — |
| SC19 | 30 | M | cough, hemoptysis, evening rise of temperature, chest pain, left pleural effusion, non- homogenous opacities in the lungs | — | .. | — |
| SC20 | 35 | M | cough, fever, weight loss, chest pain, left pleural effusion | — | — | — |
| SC21 | 58 | F | persistent cough | — | .. | — |
| SC22 | 60 | F | persistent cough, fever, right pleural effusion, multiple cavitory lesions in the left lung | — | — | — |
| SC23 | 57 | M | cough, fever, chest pain, nodular densities in the lung | — | .. | — |
| SC24 | 40 | F | evening rise of temperature accompanied with chills, cough, pulmonary infiltrates in the left lung | — | .. | — |
| SC25 | 45 | M | cough, weight loss, hemoptysis, chills, non-homogenous opacities in the lungs | — | — | — |
| SC26 | 68 | M | productive cough, weight loss, lassitude, cavitory lesion in right middle and lower lobes | — | .. | — |
| SC27 | 39 | M | cough, intermittent fever, chest pain | — | .. | — |
| SC28 | 51 | M | severe chest pain, fever, cough, left pleural effusion | — | — | — |
| SC29 | 26 | M | cough, throat pain, loss of appetite, cavitation in right upper and middle lobes | — | .. | — |
| SC30 | 55 | M | productive cough, chest pain, lassitude, cavitation in left upper lobe | — | — | — |
| SC31 | 46 | F | cough, fever, weight loss, loss of appetite, non-homogenous opacities in the lungs | — | — | — |
| SC32 | 53 | F | cough, fever, night sweats | — | .. | — |
| SC33 | 51 | M | cough, fever, weight loss, loss of appetite, chest pain, pleural effusion in both the lungs | — | .. | — |

.. Test was not performed

**Table E5.**
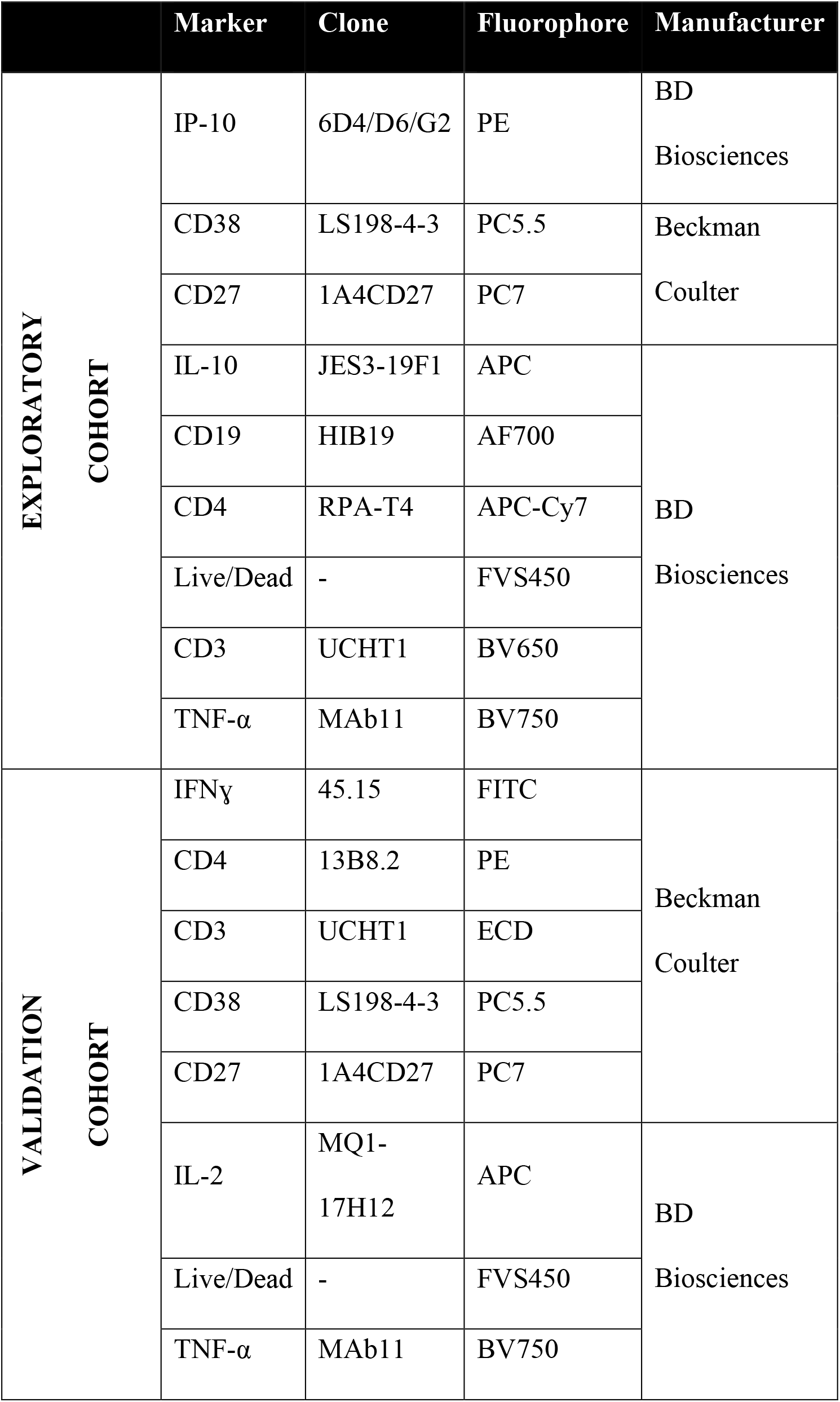
Summary of the antibody panels used in the study.

**Table E6.**
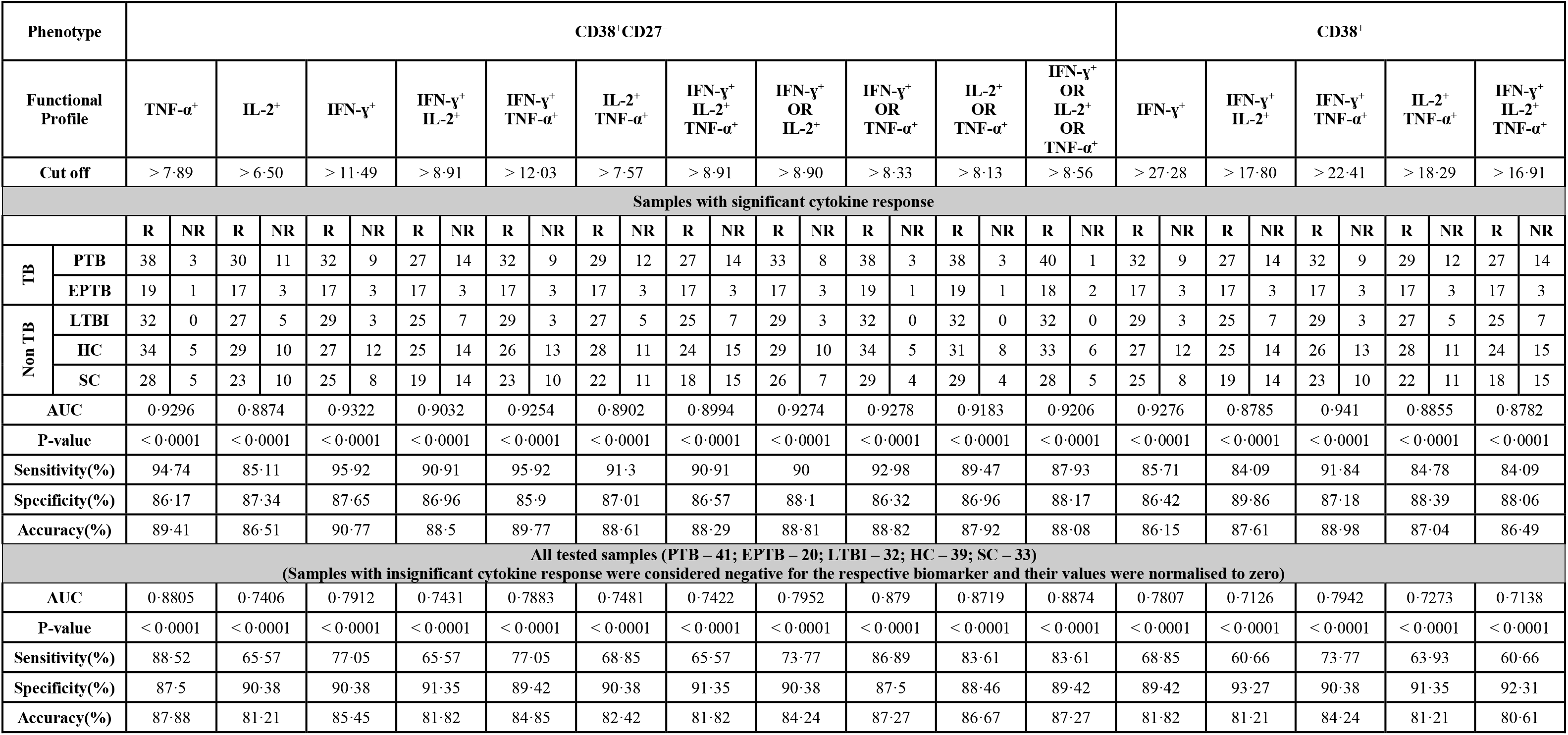
Biomarker performance of CD38^+^CD27^−^ and CD38^+^ within mono- and polyfunctional CD4^+^ T cells. Diagnostic potentials of several biomarkers with AUC values > 0.875 in *Mtb*-CW stimulated whole blood have been summarised. Number of samples with significant responses for each cytokine or their combination have been listed. For AND combinations, response was considered significant if all the cytokines involved in the combination were independently significant. For OR combinations, criteria for significant responses were same as that for TNF-α. To counter the problem of insignificant cytokine responses in whole blood, OR combinations were evaluated to check if antibodies for 2 or more cytokines could be used in the same detector channel. Mann Witney *U* test for used to compare the TB (PTB and EPTB) and non-TB (LTBI, HC and SC) groups. AUC: area under the curve.

